# Sleep inertia drives the association of evening chronotype with psychiatric disorders: epidemiological and genetic evidence

**DOI:** 10.1101/2024.09.10.24313197

**Authors:** Angus C. Burns, Stephanie Zellers, Daniel P. Windred, Iyas Daghlas, Nasa Sinnott-Armstrong, Martin Rutter, Christer Hublin, Eleni Friligkou, Renato Polimanti, Andrew J. K. Phillips, Sean W. Cain, Jaakko Kaprio, Hanna M. Ollila, Richa Saxena, Jacqueline M. Lane

## Abstract

Evening chronotypes (a.k.a. “night-owls”) are held to be at greater risk for psychiatric disorders. This is postulated to be due to delayed circadian timing increasing the likelihood of circadian misalignment in an early-oriented society. Circadian misalignment is known to heighten sleep inertia, the difficulty transitioning from sleep to wake characterized by low arousal and cognitive impairment, and evening chronotypes experience greater sleep inertia. Therefore, difficulty awakening may explain the relationship between evening chronotype and psychiatric disorders by acting as a biomarker of circadian misalignment. In analyzing the longitudinal incidence of psychiatric disorders in the UK Biobank (*n* = 496,820), we found that evening chronotype predicted increased incidence of major depressive disorder, schizophrenia, generalized anxiety disorder and bipolar disorder. Crucially, this effect was dependent on sleep inertia, which was a much stronger predictor of these disorders, such that evening types without sleep inertia were at no higher risk as compared to morning types. Longitudinal analyses of suicide and depressed mood (CES-D score) in the Older Finnish Twin Cohort (*n* = 23,854) replicated this pattern of results. Twin and genome-wide association analyses of difficulty awakening identified the trait to be heritable (Twin *H*^2^ = 0.40; SNP *h*^2^ = 0.08), enriched for circadian rhythms genes and have substantial shared genetic architecture with chronotype. Marginal and conditional Mendelian randomization analyses mirrored the epidemiological results, such that the causal effect of evening chronotype on psychiatric disorders was driven by shared genetic architecture with difficulty awakening. In contrast, difficult awakening was strongly causally associated with psychiatric disorders independently of chronotype. Psychiatric disorders were only weakly reverse causally linked to difficult awakening. Collectively, these results challenge the notion that evening chronotype is a risk factor for psychiatric disorders *per se*, suggesting instead that evening types are at greater risk for psychiatric disorders due to circadian misalignment, for which sleep inertia may be acting as a biomarker.

## Introduction

Evening chronotypes are at increased risk for mood disturbance and psychiatric disorders, including depression, anxiety, schizophrenia and bipolar disorder [1–8]. The timing of the intrinsic circadian pacemaker is substantially delayed in evening chronotypes as compared to morning chronotypes, with melatonin and core body temperature rhythms occurring on average 2-3 hours later [9, 10]. The interaction of delayed circadian phase with an early-oriented society produces misalignment between the intrinsic pacemaker and sleep-wake, activity and environmental rhythms in some individuals [11–13]. Circadian misalignment is trans-diagnostically observed in psychiatric disorders and precedes the onset of symptoms in mood disorders [14–16]. Therefore, it has been proposed that the increased risk for circadian misalignment among evening chronotypes drives the association with psychiatric disorders, rather than delayed timing of the circadian clock *per se* [2, 17].

Sleep inertia is the transition state between sleep and wakefulness that is defined by transient cognitive impairment, low arousal and difficulty awakening and is a distinct component of the human arousal state to the circadian and homeostatic processes [18–21]. The magnitude of sleep inertia is strongly influenced by circadian phase, such that waking closer to one’s core body temperature minimum (i.e., misaligned wake in the biological night) produces worse sleep inertia [22, 23]. This effect is independent of, but exacerbated by, prior sleep loss [24, 25]. Evening chronotypes experience greater sleep inertia, due to a greater likelihood of waking in the biological night and this phenomenon is most pronounced on workdays [26–28]. Therefore, it is plausible that sleep inertia drives the association of evening chronotype with psychiatric disorders by acting as a biomarker of misaligned wake and that evening types without sleep inertia are not at increased risk for developing these disorders. As yet, no study has tested this hypothesis.

Here, we investigated whether sleep inertia explained the increased incidence of psychiatric disorders and suicide among evening chronotypes in longitudinal analyses of two large, independent cohorts (UK Biobank & Older Finnish Twin Cohort). We then completed twin and genome-wide association analyses of sleep inertia to examine its heritability and shared polygenicity with chronotype, other sleep traits and psychiatric disorders. Finally, we conducted marginal and conditional Mendelian randomization analyses to examine whether the genetically instrumented causal effect of evening chronotype on psychiatric disorders is explained by an underlying shared genetic architecture with sleep inertia.

## Methods

### Participants

The cross-sectional and longitudinal analyses of psychiatric disorders and the genome-wide association study of difficulty awakening drew on the UK Biobank prospective general population cohort, which contains more than 502,000 UK residents recruited via National Health Service patient registers from 2006 to 2010. The study population is described in detail elsewhere [29, 30]. Accelerometry and light data were measured in a subset of 103,720 participants in 2013–2015 [31, 32], and a separate subset of 157,366 participants completed an online mental health questionnaire (MHQ) in 2016–2017 [33]. Participants who accepted the invitation to join the UK Biobank cohort provided written, informed consent and were given reimbursement for travel expenses. The UK Biobank has received ethical approval from the North West Multi-Center Research Ethics Committee (ref 11/NW/03820).

The longitudinal analysis of suicide and the twin-modeling analyses of difficulty awakening and chronotype drew on the Older Finnish Twin Cohort (OFTC) which is a long-running prospective cohort study that started in 1975 by mailing a baseline questionnaire to all Finnish same-sex twin pairs that were born before 1958 and where both co-twins were alive in 1975 (89% response rate) [34]. Briefly, twin pairs were selected from the Central Population Registry of Finland in 1974, and twin zygosity was determined by a validated questionnaire shown to accurately classify 93% of twin pairs as monozygotic (MZ) or dizygotic (DZ) [35]. Three follow-up health surveys were conducted, mailed to all participants in 1981, and to twins born between 1930 and 1957 in 1990 and to twins born 1945-1957 in 2011.

### Measurement of Predictors

In the UK Biobank cohort, sleep inertia was measured by self-reported difficulty awakening (data-field 1170) and chronotype was measured by self-reported diurnal preference (data-field 1180) as part of the baseline questionnaire between 2006 to 2010. Both items were drawn from the Horne-Ostberg Morningness-Eveningness Questionnaire [10] and were binarized as follows for analysis: difficulty awakening (0 = “Very easy” or “Fairly easy” *vs.* 1 = “Not very easy” or “Not at all easy”) and chronotype (0 = “Definitely a morning person” or “More a ‘morning’ than ‘evening’ person” *vs.* 1 = “More an ‘evening’ than a ‘morning” person” or “Definitely an evening person”). The OFTC study measured difficulty awakening with an item on the time required to feel fully alert after sleep and measured chronotype with an item on diurnal preference, both taken from the Diurnal Type Scale in the 1981 questionnaire [36]. These items were binarized as follows for analysis: difficulty awakening (0 = “less than 10 minutes” *vs.* 1 = “10-19 minutes” or “20-39 minutes” or “more than 40 minutes”) and chronotype (0 = “I am clearly a morning person” or “I am to some extent a morning person” *vs.* 1 = “I am to some extent an evening person” or “I am clearly an evening person”; See Supplementary Methods for more details).

### Measurement of Outcomes

For the UK Biobank light and sleep analyses, we used data from 103,720 participants who completed objective monitoring of physical activity, sleep and light across 7 continuous days. Participants wore an AX3 triaxial accelerometer (Axivity, Newcastle upon Tyne, UK) with in-built light sensor (APDS9007 silicon photodiode sensor; spectral sensitivity λ = 470-650nm) on their dominant wrist continuously for 7 days, while continuing with their normal activities. The analysis of light and sleep data was completed in R (version 4.1.0) and followed previously reported methodology [31] (see Supplementary Methods for a detailed description). The R packages *GGIR* (v1.6.9) and *sleepreg* (v1.3.4) [32, 37, 38] were used to assess accelerometer data quality and provide summaries of device non-wear and extract sleep parameters. Sleep duration, sleep efficiency, sleep midpoint timing and the sleep regularity index (SRI; a measure of day-to-day intra-individual sleep-wake variability) were derived [38, 39]. Day and night light exposure values (day, 7.30am-8.30pm; night, 12.30am-6am) were ascertained from individual light exposure profiles according to previously published methods [31].

For the UK Biobank longitudinal analyses, International Classification of Diseases, Tenth Revision, Clinical Modification (ICD10) codes were used to determine the incidence of major depressive disorder (UK Biobank F code, F32), schizophrenia (F20), generalized anxiety disorder (F41) and bipolar disorder (F31) and death registry data was used to confirm the vital status of participants (data-field 40000).

For the UK Biobank cross-sectional analyses, the responses of 157,366 participants who completed the Mental Health Questionnaire were used. The definitions of psychiatric disorder outcomes from the MHQ are based on Composite International Diagnostic Interview (CIDI) and Diagnostic and Statistical Manual IV (DSM-IV) criteria and followed guidelines established by Davis, Coleman [33] and reported previously [31]. For the UK Biobank cross-sectional analyses, psychiatric outcomes were major depressive disorder (MDD), generalized anxiety disorder (GAD), bipolar disorder (hypomania/mania), post-traumatic stress disorder (PTSD), psychotic experiences, and self-harm derived from the MHQ. Detailed definitions are given in the Supplementary Methods.

In the Older Finnish Twin Cohort longitudinal analyses depressive symptoms were measured by the Center for Epidemiologic Studies Depression (CES-D) total score from the 2011 survey [40].Vital status (alive in Finland on December 31, 2020, date of death or date of migration from Finland) as well as date of birth and sex provided at the formation of the cohort) were obtained from the Population Information System (dvv.fi), Finland. Cause of death data was provided by Statistics Finland and deaths attributed to suicide were extracted as an outcome.

### Statistical Analyses

The longitudinal association of difficulty awakening and chronotype with risk for incident psychiatric disorders in the UK Biobank was examined using Cox proportional hazard regression models: two marginal models (difficulty awakening alone; chronotype alone) and one conditional model (difficulty awakening and chronotype together). Each of these models was tested in two nested models with increasing adjustment for potential confounders. Model 1 adjusted for age, sex and season of assessment. Model 2 additionally adjusted for physical activity and employment status (employed *vs.* unemployed; data-field 6142). Physical activity was measured using the short International Physical Activity Questionnaire (IPAQ) and metabolic equivalent (MET) minutes of physical activity per week were calculated according to published guidelines [41]. Individuals diagnosed with the relevant disorder prior to the baseline questionnaire were excluded and for non-diagnosed individuals time-to-event data were right-censored at the time of participant mortality or the study end date. Hazard ratios and their 95% confidence intervals are reported. The proportional hazards assumption was assessed for each model. A sensitivity analysis examined the longitudinal associations in the UK Biobank cohort after additionally adjusting for sleep duration (data-field 1160), sleep duration-squared and daytime sleepiness (data-field 1220; longitudinal Model 3). The longitudinal analysis in the Older Finnish Twin Cohort of CES-D scores and suicide followed the same pattern of two marginal models and one conditional model. Each model was adjusted for age, sex and season of assessment. The association with CES-D score was examined with multiple linear regression, with standardized beta values and their 95% confidence intervals reported. The association with suicide was examined with Cox proportional hazards models, with hazard ratios and their 95% confidence intervals reported.

The cross-sectional associations of difficulty awakening and late chronotype with each of the psychiatric case-control, sleep and light variables in the UK Biobank were examined following the same pattern of two marginal models examining either difficulty awakening or chronotype and one conditional model examining both. Two nested models with the same covariate adjustment as in the longitudinal models were tested. The association of difficulty awakening and late chronotype with prevalent case/control outcomes were examined with multiple logistic regression. Odds ratios (OR) and their 95% confidence intervals are reported. Multiple linear regression was used for the continuous objectively measured sleep and light parameters, with standardized beta values reported. A sensitivity analysis examined the cross-sectional associations in a subset of the UK Biobank cohort that completed the objective sleep measurement (*n* = 86,772) after additionally adjusting for objective sleep duration, sleep efficiency, sleep midpoint timing and physical activity (as opposed to MET minutes; cross-sectional Model 3).

All twin analyses were conducted in R using the package OpenMx. First, we calculated within-pair correlations stratified by zygosity (MZ and DZ). Second, we evaluated the genetic and environmental variance underlying sleep inertia and chronotype, as well as their genetic and environmental covariance, via a biometric variance decomposition (twin model). Twin models leverage the amount of genetic variation shared by MZ and DZ twins; MZ twins share 100% of their genetic variation, whereas DZ twins share only 50%, the same as any other pair of full siblings. MZ and DZ twin pairs both share many environmental and other factors (e.g., birth cohort, age, prenatal conditions, socioeconomic status, neighborhood, and rearing environment) but also have unique experiences across development. These known similarities within twin pairs allow for the decomposition of trait variance into various sources: additive genetic effects (A), genetic dominance effects (D), shared environmental influences (C), and unique environmental influences (E). Shared environmental influences are those factors that make twins in a pair more similar to each other, whereas unique environmental influences are those factors that make twins in a pair more dissimilar to each other. We first estimated a univariate twin model for sleep inertia to determine which variance components best explained the observed familial similarity (A, C, D, and/or E). Univariate results in the same cohort have already been published for chronotype [42]. We then estimated a bivariate twin model of sleep inertia and chronotype with the most appropriate variance components included for each phenotype based on the univariate results. Reporting of statistical analyses and results followed the RECORD guidelines.

### Genome-wide Association Studies of Difficulty Awakening and Chronotype

We performed the genetic association tests for difficulty awakening using REGENIE version 3.0.3 [43], controlling for age, sex, genotyping array (UKBiLEVE array *vs*. UKB Axiom) and the top 10 principal components of ancestry in 457,737 individuals of European ancestry (see Supplementary Methods for details on ancestry definition). REGENIE is a two-step whole-genome regression approach that accounts for potential population stratification and sample relatedness allowing us to include the full sample of related European participants in the UK Biobank. Specifically, in step 1 of REGENIE, we used a subset of 156,817 LD-pruned (*r*^2^ < 0.10, window = 500kb) SNPs with MAF >1% to calculate a leave-one-chromosome-out (LOCO) polygenic score for each trait and each individual using ridge regression. Association testing was then performed, in step 2 of REGENIE, including the LOCO polygenic predicted value from step 1 as an offset in the linear regression model in addition to other covariates, to account for sample relatedness for 18,934,942 variants (MAF > 0.001, INFO > 0.30). Independent genomic risk loci reaching genome-wide significance (*p* < 5×10^−8^) were defined as those in low linkage-disequilibrium (*r*^2^ < 0.10) within a 250kb block. For chronotype, we used the previously published inverse-variance weighted meta-analysis of chronotype from the UK Biobank and 23andMe samples for 15,880,664 variants in up to 651,295 individuals [44].

### Post-GWAS Analyses

We used genome-wide complex trait Bayesian analysis (GCTB, SbayesS, v2.0) to estimate the SNP-based heritability (SNP *h*^2^), polygenicity and selection coefficients for difficulty awakening and chronotype [45]. Univariate linkage disequilibrium score regression (LDSR) was also used to estimate SNP-based heritability and the LDSR intercept, which captures the degree of population stratification [46]. We then used bivariate Gaussian mixture modeling implemented in MiXeR (v1.3) to examine the degree and directional consistency of the shared genetic architecture between difficulty awakening and chronotype (see Supplementary Methods for details). MAGMA was used to assess the enrichment of difficulty awakening loci in genes, molecular pathways and tissues as implemented in FUMA with the default parameter settings [47, 48].

To assess the genetic correlation of difficulty awakening and chronotype with sleep and psychiatric outcome traits we applied bivariate LDSR using the latest GWAS for each outcome (see Supplementary Methods for external GWAS meta-data). The sleep traits included were accelerometry-derived mid-sleep point, most-active 10h timing, sleep duration and sleep efficiency as well as self-reported insomnia symptoms, daytime sleepiness, and napping behaviour [49–52]. The psychiatric disorder traits included major depressive disorder, attempted suicide, schizophrenia, PTSD, anxiety disorders and bipolar disorder [53–58].

### Conditional GWAS & Mendelian Randomization Analyses

We used multi-trait conditional and joint analysis (mtCOJO) and generalized summary-data-based Mendelian randomization (GSMR) to conduct marginal and conditional MR analyses of difficulty awakening and chronotype as exposures on sleep-wake timing and psychiatric outcome traits [59]. The mtCOJO method provides genome-wide conditioning of the exposure GWAS SNP effects (e.g. difficulty awakening) on covariate GWAS SNP effects (e.g. chronotype) to identify genetic variants for the exposure that are independent of the covariate. This approach produces conditional SNP effects that are free from collider bias which can arise when using the conditioned variable as a covariate in the initial GWAS and is robust to sample overlap [60]. The GSMR method improves on previous MR methods by modeling the error in the SNP-exposure estimate and removing SNPs with horizontal pleiotropic effects using the heterogeneity in dependent instrument (HEIDI) outlier test, thus boosting power and reducing type-1 error [59]. The GSMR framework also allows for the conditional analysis of the exposure-outcome causal association by first completing mtCOJO to identify genetic variants for the exposure that are independent of the conditioned covariate (see Supplementary Methods for details). Furthermore, GSMR is well-calibrated to detect independent forward (exposure to outcome) and reverse (outcome to exposure) causal effects, including discordant effects [59]. Here, we completed a series of marginal and conditional MR analyses for difficulty awakening and chronotype before and after mutual mtCOJO adjustment on sleep-wake timing and psychiatric outcome traits.

## Results

### Participant characteristics of UK Biobank and Older Finnish Twin Cohort samples

The participant characteristics of the UK Biobank and OFTC samples and their missingness are presented in Supplementary Tables 1 & 2 across difficulty awakening and chronotype categories. In both samples, the phenotypic correlation between sleep inertia phenotypes and chronotype was moderate (UK Biobank, *r* = 0.42, *p* < 0.0001; OFTC, *r* = 0.44, *p* < 0.0001). In the UK Biobank sample, evening types had seven-fold higher odds of reporting difficult awakening (odds ratio OR = 7.02, 95% CI = 6.89 – 7.15, *p* < 0.0001), as compared to morning types. Of note, this relationship held and was slightly stronger when adjusting for subjective sleep duration, daytime sleepiness and insomnia symptoms, indicating that greater risk of difficult awakening among evening types was not explained by these sleep quality-related variables (OR = 7.35, 95% CI = 7.22 – 7.48, *p* < 0.0001).

### Sleep inertia and chronotype are associated with objectively measured sleep and light exposure parameters

We first examined the association of difficulty awakening and chronotype with objectively measured sleep and light exposure parameters in a set of marginal and conditional models within the subset of the UK Biobank cohort that completed the seven-day actigraphy study (*n* = 86,772; 5.7 ± 1.1 years after the baseline questionnaire; Supplementary Table 3). In the fully adjusted conditional models (Model 2), evening chronotype was associated with later mid-sleep timing but was not associated with lower Sleep Regularity Index (SRI) score independent of difficulty awakening (mid-sleep timing β = 0.45, 95% CI = 0.44 0.47, *p* < 0.0001; SRI β = –0.01, 95% CI = –0.03 0.002, *p* = 0.10). In contrast, difficulty awakening was independently associated with both later mid-sleep timing and reduced SRI score (mid-sleep timing β = 0.14, 95% CI = 0.12 0.16, *p* < 0.0001; SRI β = –0.05, 95% CI = –0.07 –0.03, *p* < 0.0001). In a sensitivity analysis that additionally adjusted for objectively measured sleep duration and sleep efficiency evening chronotype remained associated with later mid-sleep timing and not with SRI (mid-sleep timing β = 0.45, 95% CI = 0.44 0.47, *p* < 0.0001; SRI β = 0.003, 95% CI = –0.01 0.02, *p* = 0.68) and difficult awakening remained associated with both later mid-sleep timing and reduced SRI (mid-sleep timing β = 0.14, 95% CI = 0.12 0.16, *p* < 0.0001; SRI β = –0.06, 95% CI = – 0.08 –0.04, *p* < 0.0001). Evening chronotype was also associated with reduced sleep efficiency and shorter nightly sleep duration (sleep efficiency β = –0.08, 95% CI = –0.10 –0.06, *p* < 0.0001; sleep duration β = –0.07, 95% CI = –0.09 –0.06, *p* < 0.0001) while difficult awakening was associated with reduced sleep efficiency and slightly longer sleep duration (sleep efficiency β = – 0.08, 95% CI = –0.10 –0.06, *p* < 0.0001; sleep duration β = 0.03, 95% CI = 0.004 –0.05, *p* = 0.02). Finally, evening chronotype was associated with reduced daytime light exposure and increased nighttime light exposure (daytime light β = –0.04, 95% CI = –0.06 –0.03, *p* < 0.0001; nighttime light β = 0.11, 95% CI = 0.09 0.13, *p* < 0.0001), independent of difficulty awakening. As well, difficult awakening was associated with reduced daytime light exposure and increased nighttime light exposure (daytime light β = –0.03, 95% CI = –0.04 –0.01, *p* = 0.002; nighttime light β = 0.03, 95% CI = 0.01 0.05, *p* = 0.005), independent of chronotype.

### Longitudinal associations of sleep inertia and chronotype with incident psychiatric disorders in the UK Biobank

In fully adjusted marginal models (longitudinal Model 2), evening chronotype was associated with a higher risk of developing major depressive disorder (aHR = 1.19, 95% CI = 1.16-1.23, *p* < 0.0001), schizophrenia (aHR = 1.30, 95% CI = 1.06-1.58, *p* = 0.01), generalized anxiety disorder (aHR = 1.20, 95% CI = 1.17-1.24, *p* < 0.0001), and bipolar disorder (aHR = 1.37, 95% CI = 1.18-1.59, *p* = 0.0006) as compared to morning chronotypes across an average of 16.58 ± 2.06 years of follow-up (Figure 1, Supplementary Table 4). Similarly, difficult awakening was associated with higher risk of developing major depressive disorder (aHR = 1.82, 95% CI = 1.76-1.88, *p* < 0.0001), schizophrenia (aHR = 2.69, 95% CI = 2.20-3.28, *p* < 0.0001), generalized anxiety disorder (aHR = 1.77, 95% CI = 1.71-1.82, *p* < 0.0001) and bipolar disorder (aHR = 1.91, 95% CI = 1.63-2.23, *p* < 0.0001), as compared to those with easy awakening and with generally larger effect sizes than those of the chronotype marginal effects. However, in the conditional model when chronotype was conditioned on difficulty awakening there was no independent association of evening chronotype risk for developing major depressive disorder (aHR = 0.99, 95% CI = 0.96-1.03, *p* = 0.63), schizophrenia (aHR = 0.93, 95% CI = 0.74-1.16, *p* = 0.51), generalized anxiety disorder (aHR = 1.01, 95% CI = 0.98-1.04, *p* = 0.57) or bipolar disorder (aHR = 1.13, 95% CI = 0.96-1.33, *p* = 0.15; Figure 1, Supplementary Table 4). In contrast, when difficulty awakening was conditioned on chronotype there remained a strong independent association of difficult awakening with increased risk for developing major depressive disorder (aHR = 1.81, 95% CI = 1.74-1.88, *p* < 0.0001), schizophrenia (aHR = 2.68, 95% CI = 2.12-3.38, *p* < 0.0001), generalized anxiety disorder (aHR = 1.73, 95% CI = 1.67-1.79, *p* < 0.0001) and bipolar disorder (aHR = 1.84, 95% CI = 1.53-2.20, *p* < 0.0001).

**Figure 1.**
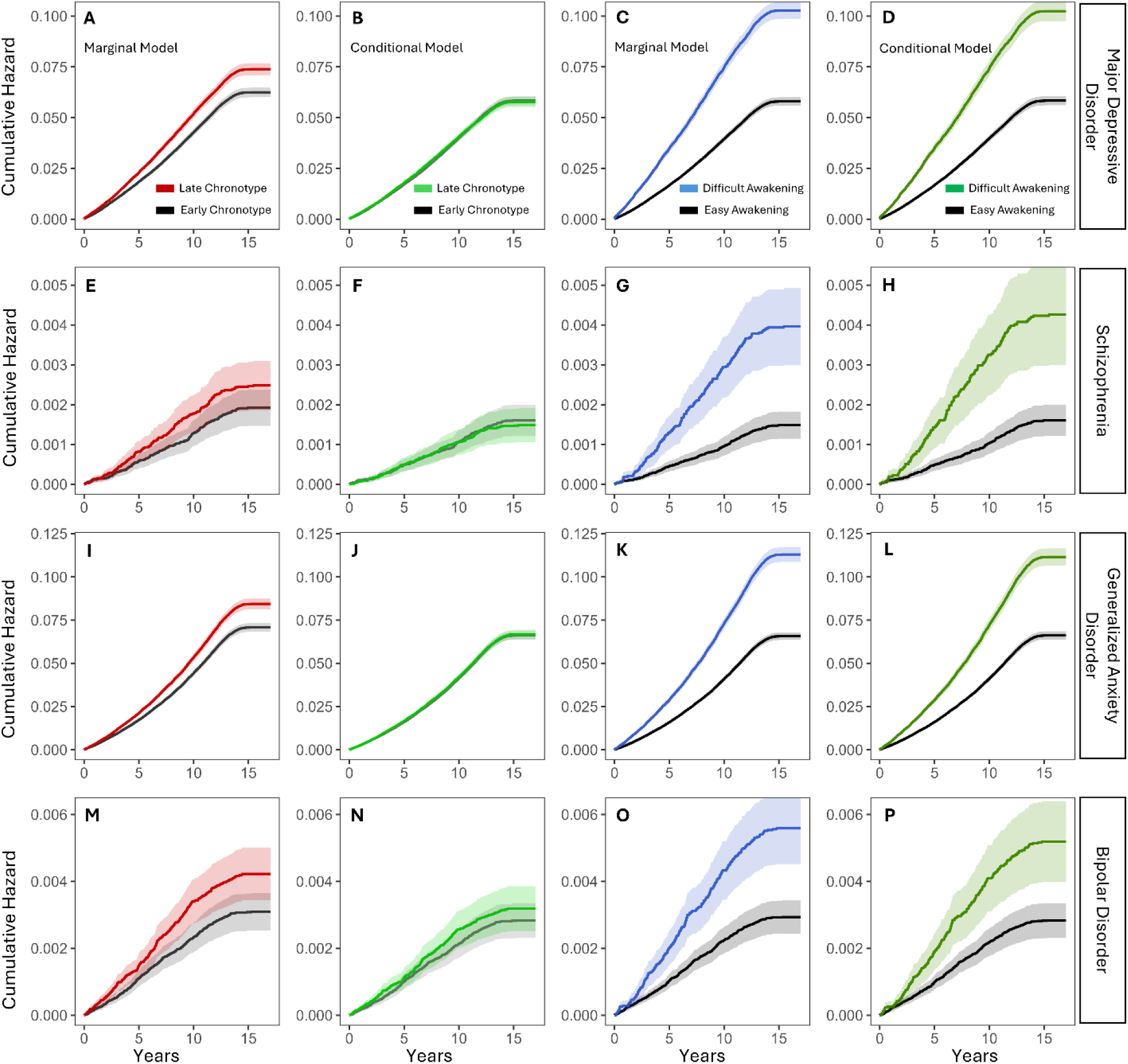
Cumulative hazard plots for the marginal and conditional longitudinal associations of difficult awakenin and evening chronotype with (A-D) major depressive disorder, (E-H) schizophrenia, (I-L) generalized anxiet disorder and (M-P) bipolar disorder. The cumulative incidence ± the 95% CI for difficult awakening *vs.* easy awakening and late chronotype *vs.* early chronotype in marginal and conditional models are presented. Model 2 is presented adjusted for age, sex, ethnicity, season, employment, and physical activity. Cumulative incidence for the marginal models are displayed in blue (difficult awakening) and red (late chronotype) and cumulative incidence for the conditional model of difficult awakening and late chronotype after mutual adjustment is displayed in green. In marginal models, difficulty awakening and chronotype are included separately, whereas in conditional models difficulty awakening and chronotype are included together Note: flattening at the time of maximum follow-up is due to the UKB recruitment period occurring over four years.

These results were robust to additional adjustment for baseline subjective sleep duration, sleep duration-squared and daytime sleepiness, excepting small reductions in effect size (longitudinal Model 3; Supplementary Table 5).

### Cross-sectional associations of sleep inertia and chronotype with psychiatric disorders and self-harm in the UK Biobank

In cross-sectional marginal analyses in the UK Biobank sample, difficulty awakening and evening chronotype were both associated with increased risk for major depressive disorder, bipolar disorder, generalized anxiety disorder, PTSD, psychotic experiences, and self-harm in fully adjusted marginal models (cross-sectional Model 2, Supplementary Table 6; Supplementary Figure 1) with a generally larger effect size for difficult awakening (OR range = 1.60-2.99) as compared to evening chronotype (OR range = 1.20-1.42). However, when chronotype was conditioned on difficulty awakening the independent association of evening chronotype with all outcomes was dramatically reduced and, in some cases, reversed (Supplementary Table 6; Supplementary Figure 1). Evening chronotype was still associated with a slightly increased risk of major depressive disorder after adjustment for difficulty awakening (OR = 1.05, 95% CI = 1.02-1.08, *p* = 0.001), however, the associations of evening chronotype with self-harm (OR = 1.02, 95% CI = 0.97-1.09, *p* = 0.35), bipolar disorder (OR = 1.06, 95% CI = 0.97-1.15, *p* = 0.18) and psychotic experiences (OR = 1.04, 95% CI = 0.98-1.09, *p* = 0.20) were non-significant. Remarkably, evening chronotype was associated with reduced risk for PTSD (OR = 0.95, 95% CI = 0.89-0.99, *p* = 0.03) and generalized anxiety disorder (OR = 0.92, 95% CI = 0.88-0.97, *p* = 0.004) after conditioning on difficulty awakening, such that evening chronotypes without difficulty awakening had lower risk for these disorders. In contrast, difficulty awakening remained strongly associated with risk for all psychiatric outcomes in the UK Biobank after conditioning on chronotype (Supplementary Table 6; Supplementary Figure 1). Greater difficulty awakening was associated with increased risk for major depressive disorder (OR = 2.84, 95% CI = 2.75-2.95, *p* < 0.0001), self-harm (OR = 1.63, 95% CI = 1.52-1.73, *p* < 0.0001), generalized anxiety disorder (OR = 2.44, 95% CI = 2.29-2.58, *p* < 0.0001), PTSD (OR = 2.32, 95% CI = 2.19-2.45, *p* < 0.0001), bipolar disorder (OR = 2.07, 95% CI = 1.89-2.28, *p* < 0.0001) and psychotic experiences (OR = 1.56, 95% CI = 1.46-1.66, *p* < 0.0001), such that difficulty awakening was associated with increased risk for these disorders independent of chronotype.

We replicated these associations in a sensitivity analysis examining the subset of the UK Biobank cohort that completed the objective sleep measurement (*n* = 86,772). This pattern of associations was unchanged after additional adjustment for objective sleep duration, sleep efficiency, sleep midpoint timing and physical activity (Cross-Sectional Model 3), excepting slight variations in effect size (Supplementary Table 7).

### Sleep inertia longitudinally predicts depressed mood and suicide in the Older Finnish Twin Cohort

In the Older Finnish Twin Cohort study, we replicated the pattern of associations observed in the UK Biobank sample in a longitudinal design with depressive symptoms and death by suicide as outcomes (Figure 2). In marginal models, both evening chronotype (B = 0.77, 95% CI = 0.40 1.16, *p* < 0.0001) and greater difficulty awakening (B = 2.10, 95% CI = 1.71 2.49, *p* < 0.0001) were associated with increased depressed mood symptoms measured by CES-D score. However, in the conditional model only difficult awakening remained longitudinally associated with depressive symptoms after conditioning on chronotype (B = 2.05, 95% CI = 1.63 2.46, *p* < 0.0001). Chronotype was not associated with depressive symptoms after conditioning on difficulty awakening (B = 0.14, 95% CI = –0.26 0.53, *p* = 0.49). In marginal models, difficult awakening was associated with increased hazard of suicide (aHR = 1.53, 95% CI = 1.17 2.00, *p* = 0.002) and there was directionally consistent but non-significant association with evening chronotype (aHR = 1.19, 95% CI = 0.91 1.56, *p* = 0.21). The association of difficulty awakening with increased risk of suicide remained after conditioning on chronotype (aHR = 1.51, 95% CI = 1.14 2.00, *p* = 0.004), while the hazard ratio for chronotype was reduced after conditioning on difficulty awakening (aHR = 1.06, 95% CI = 0.80 1.41, *p* = 0.43, Figure 2).

**Figure 2.**
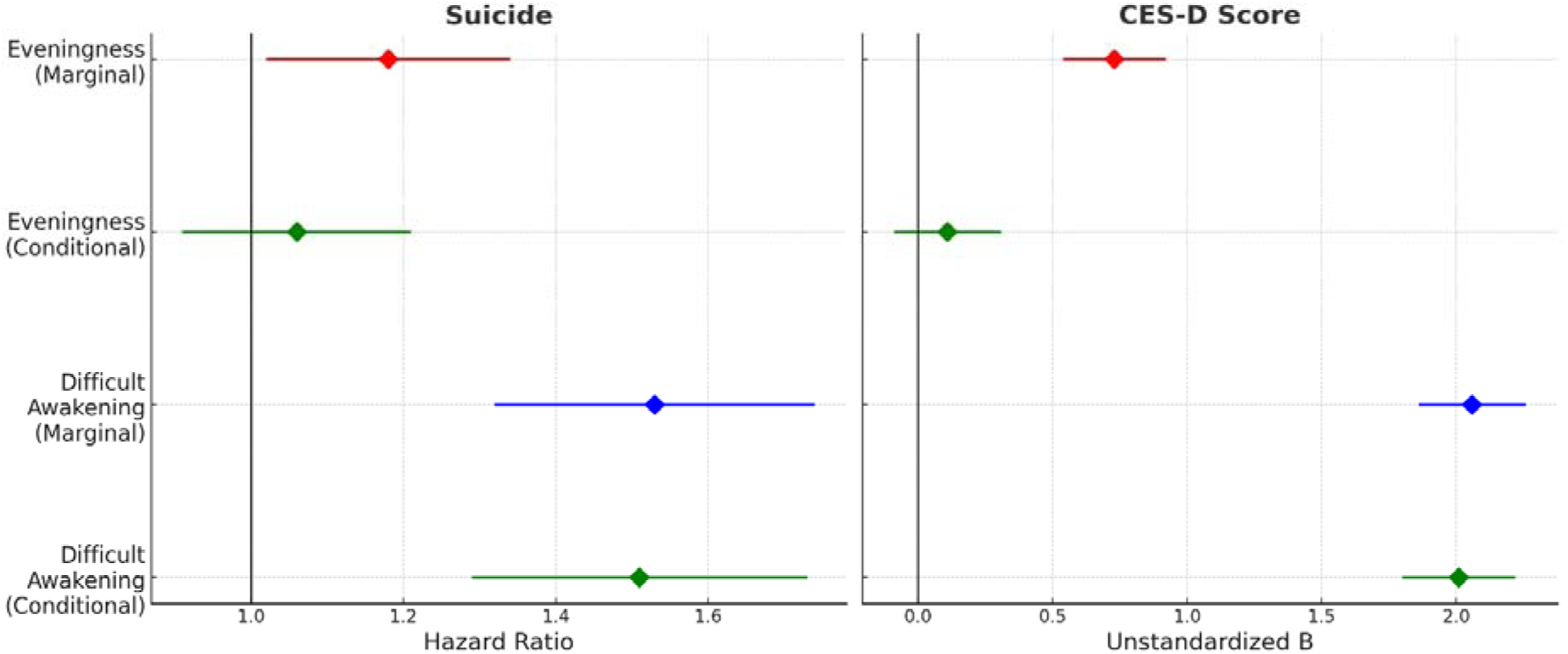
Coefficients plot for the longitudinal marginal and conditional association of difficult awakening and late chronotype with (A) suicide and (B) Center for Epidemiologic Studies Depression (CES-D) score in the Older Finnish Older Twin Cohort study (*n* = 23,095). Coefficients represent hazard ratios ± SEM for (A) suicide and unstandardized b coefficients ± SEM for (B) CES-D score.

### Twin analyses of sleep inertia and chronotype demonstrate heritability, shared genetic and environmental influences

Twin analyses completed using the OFTC sample demonstrated the heritability of difficulty awakening and chronotype as well as their shared genetic and environmental influences. Within-pair correlations for both sleep inertia and chronotype were larger in MZ twins (sleep inertia correlation = 0.40, *p* < 0.0001; chronotype correlation = 0.48, *p* = < 0.0001) as opposed to DZ twins (sleep inertia correlation = 0.16, *p* < 0.0001; chronotype correlation = 0.15, *p* < 0.0001), indicating genetic influences on both phenotypes.

Univariate model results for sleep inertia indicated that an ADE model was most appropriate. The published results for chronotype (Koskenvuo et al., 2006) also indicated an ADE model. Therefore, the final model was a bivariate model with ADE components for both phenotypes and their covariance. Both phenotypes were heritable (*H*^2^ inertia = 0.40, 95% CI = 0.35-0.45; *H*^2^ chronotype = 0.48, 95% CI = 0.45-0.51) with additive genetic (difficulty awakening *A* = 0.25; chronotype *A* = 0.10) and dominance effects (difficulty awakening *D* = 0.15; chronotype *D* = 0.38). There was also significant environmental influence over both traits (difficulty awakening *E* = 0.60; chronotype *E* = 0.52). The additive and dominance genetic correlations were strongly positive (*r_A_* = 0.83, *p* = 0.01; *r_D_* = 0.64, *p* = 0.001). There was also a modest environmental correlation (*r_E_* = 0.27, *p* < 0.0001).

### Genome-wide association study of difficulty awakening

Given that we observed significant broad-sense heritability of difficulty awakening and genetic correlations with chronotype in the OFTC twin analyses, we undertook a genome-wide association study (GWAS) of difficulty awakening in the UK Biobank cohort in 457,737 individuals of European ancestry to further investigate this relationship. The GWAS of difficulty awakening identified 98 independent genomic risk loci passing the genome-wide significance threshold (*p* < 5×10^−8^; λ_GC_= 1.52; mean χ^2^ = 1.69; LDSR Intercept = 1.04, SE = 0.01; Figure 3, Supplementary Table 8). The difficulty awakening trait was heritable (LDSR SNP *h*^2^ = 0.07, SE = 0.003; SBayesS SNP *h*^2^ = 0.08, SD = 0.001), though less so than chronotype (LDSR SNP *h*^2^ = 0.18, SE = 0.005; SBayesS SNP *h*^2^ = 0.11, SD = 0.001). The selection coefficient for both difficulty awakening (*S* = –0.61, SD = 0.04) and chronotype (*S* = –0.48, SD = 0.03) was negative, though stronger for difficulty awakening suggesting stronger purifying selection on loci for this trait (Supplementary Table 9). The estimated polygenicity of difficulty awakening (π = 0.033, SD = 0.002) was slightly higher than that of chronotype (π = 0.029, SD = 0.001).

**Figure 3.**
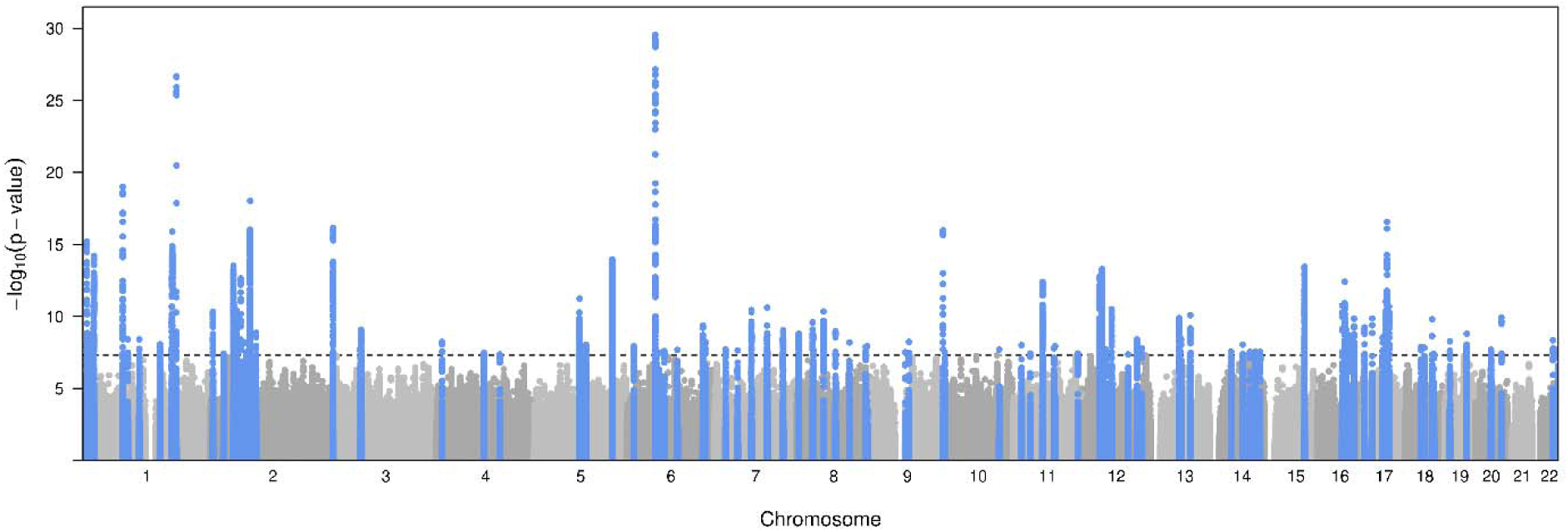
Manhattan plot for the genome-wide association study of difficulty awakening in the UK Biobank cohort. Peaks highlighted in blue represent genomic risk loci passing the genome-wide significance threshold, *p* < 5×10^−8^.

We performed bivariate Gaussian mixture modelling to assess the degree and directional consistency of the shared genetic architecture between difficulty awakening and chronotype. Difficulty awakening and chronotype were strongly genetically correlated, with the estimate mirroring the additive genetic correlation obtained in the twin models (MiXeR *r*_g_ = 0.82, SE = 0.004; Supplementary Figure 2, Supplementary Table 10). Approximately 9,100 common variants were estimated to influence difficulty awakening while 8,600 variants were estimated to influence chronotype. Of the difficulty awakening variants, 820 variants were unshared with chronotype and of the chronotype variants, 300 were unshared with difficulty awakening (i.e., ∼900 total unshared variants). When considering the number of shared variants as a proportion of the total polygenicity of difficulty awakening and chronotype, approximately 93% of the variants associated with difficulty awakening were found to also be associated with chronotype (dice coefficient = 0.93, SD = 0.02). However, there was significant discordance among these variants, with approximately 16% (SE = 1%) of the shared variants showing opposing directions of effect (Supplementary Figure 2, Supplementary Table 10).

MAGMA gene enrichment tests identified 185 Bonferroni-significant genes enriched for difficulty awakening loci, of which 68 overlapped with significantly enriched genes from chronotype MAGMA gene-based tests (see Supplementary Tables 11 and 12). These included known circadian rhythms genes, including *PER2, PER3, CRY1, CIART, FBXL3* as well as genes involved in ancillary pathways such as G protein-coupled receptor signaling (*NMUR2, RGS16, GPR61*), mitogen activated protein kinase signaling (*RASA4, RASD1*, *MAPK12*) and synaptic organization (*NRXN1, SHANK2*). Pathway enrichment analyses found Bonferroni-significant enrichment in the synaptic membrane adhesion, synapse assembly and synapse organization pathways (Supplementary Table 13). Tissue enrichment analyses found genes expressed in the brain and pituitary gland were enriched for difficulty awakening loci, as well as genes expressed in several subordinate brain regions including the hypothalamus, however there was no difference in enrichment across brain developmental stages (Supplementary Tables 14 and 15, Supplementary Figure 3).

### Genetic overlap of difficulty awakening and chronotype with sleep-wake and psychiatric traits

We next used bivariate linkage disequilibrium score regression (LDSR) to calculate and compare the genetic correlations of difficulty awakening and chronotype with sleep and psychiatric traits using the most-recent GWAS for each trait. With respect to the sleep-wake traits, both difficulty awakening and chronotype are strongly positively genetically correlated with later accelerometry-derived sleep midpoint timing (difficulty awakening *r*_g_ = 0.59, *p* = 1.63×10^−43^; chronotype *r*_g_ = 0.83, *p* = 1.37×10^−89^) and timing of the most active 10h of the day (difficulty awakening *r*_g_ = 0.67, *p* = 7.30×10^−44^; chronotype *r*_g_ = 0.84, *p* = 4.35×10^−78^). In contrast, both traits show weak genetic correlations with other sleep traits including sleep duration, sleep efficiency, insomnia symptoms and daytime sleepiness (Supplementary Table 16, Supplementary Figure 4). Of note, while late chronotype is not significantly genetically correlated with sleep duration or insomnia symptoms (*p*_both_ > 0.70), difficulty awakening shows a small genetic correlation with both longer sleep duration (*r*_g_ = 0.09, *p* = 0.002) and increased insomnia symptoms (*r*_g_ = 0.12, *p* = 4.49×10^−7^). With respect to the psychiatric traits, genetic correlations for difficulty awakening and chronotype were positive, though with larger effect sizes for difficulty awakening (*r*_g_ range = 0.11 – 0.27) compared to chronotype (*r*_g_ range = 0.03 – 0.12; Supplementary Table 16, Supplementary Figure 5).

### Conditional Genome-Wide Association Study of Difficulty Awakening and Chronotype

We next sought to complete a conditional genome-wide association study of difficulty awakening and chronotype using multi-trait Conditional and Joint Analysis (mtCOJO) to identify difficulty awakening SNP effects that are independent of chronotype, and vice versa. As expected, given the large degree of pleiotropy between difficulty awakening and chronotype, conditional analysis reduced the number of genome-wide significant loci for both traits. We identified 24 genome-wide significant genomic risk-loci for difficulty awakening that were independent of chronotype (Supplementary Table 17) and 40 genomic risk loci for chronotype that were independent of difficulty awakening (*p* < 5×10^−8^; Supplementary Table 18, Supplementary Figure 6). Eleven of these loci were shared between the conditional difficulty awakening and chronotype GWAS due to discordant SNP effects. Of note, rs4757137 in the promoter region of the core clock gene *BMAL1* was associated with increased difficulty awakening after conditioning on chronotype (β Conditional = 0.012, SE = 0.002, *p* = 2.02×10^−10^) and morning chronotype after conditioning on difficulty awakening (β Conditional = –0.05, SE = 0.007, *p* = 1.10×10^−12^). However, the marginal effect of rs4757137 on difficulty awakening was not significant (β Marginal = 0.003, SE = 0.002, *p* = 0.05), while the marginal effect on chronotype was significant (β Marginal = –0.04, SE = 0.007, *p* = 4.96×10^−9^). In contrast, the rs6968865 SNP within the *AHR* locus was only nominally associated with increased difficulty awakening (β Marginal = 0.007, SE = 0.002, *p* = 4.55×10^−5^) and morning chronotype (β Marginal = –0.02, SE = 0.006, *p* = 0.0002) in the marginal analysis, while in the conditional analysis it showed a discordant genome-wide significant association with increased difficulty awakening (β Conditional = 0.011, SE = 0.002, *p* = 3.20×10^−11^) and morning chronotype (β Conditional = – 0.04, SE = 0.006, *p* = 2.40×10^−10^) in the conditional analysis. Finally, rs16872808 in the *ZFPM2* locus was only nominally associated with reduced difficulty awakening (β Marginal = –0.005, SE = 0.002, *p* = 0.0010) and evening chronotype (β Marginal = 0.022, SE = 0.006, *p* = 0.0004) in the marginal analysis, whereas it was significantly associated with reduced difficulty awakening after conditioning on chronotype (β Conditional = –0.01, SE = 0.002, *p* = 5.44×10^−9^) and it was significantly associated with evening chronotype after conditioning on difficulty awakening (β Conditional = 0.04, SE = 0.006, *p* = 1.12×10^−8^).

### Marginal and Conditional Two-Sample Mendelian Randomization

We next examined the potential causal effects of difficulty awakening and evening chronotype on sleep-wake timing traits, psychiatric disorders and attempted suicide using Mendelian randomization (GSMR) in a set of marginal (unadjusted GWAS) and conditional models (mtCOJO conditional GWAS). We first examined whether difficulty awakening and chronotype were causally associated with accelerometry-derived mid-sleep timing and timing of the most active 10 hours of the day to extend prior MR work and the UK Biobank epidemiological findings reported here [8]. In the marginal model, genetically instrumented evening chronotype was causally associated with later sleep midpoint timing (β_xy_ Marginal = 0.28, SE = 0.009, *p* = 1.33×10^−196^) and later timing of the most active 10 hours of the day (β_xy_ Marginal = 0.27, SE = 0.009, *p* = 2.77×10^−194^; Supplementary Table 19). Critically, in the conditional model genetically instrumented evening chronotype that is independent of difficulty awakening remained associated with later mid-sleep timing (β_xy_ Conditional = 0.16, SE = 0.03, *p* = 1.05×10^−07^) and later timing of the most active 10h of the day (β_xy_ Conditional = 0.14, SE = 0.03, *p* = 1.77×10^−06^). This established that the chronotype conditional GWAS indexed later timing that is independent of difficulty awakening. In the marginal models, increased difficulty awakening was associated with later mid-sleep timing (β_xy_ Marginal = 0.81, SE = 0.05, *p* = 2.35×10^−61^) and later timing of the most active 10 hours of the day (β_xy_ Marginal = 0.82, SE = 0.05, *p* = 5.15×10^−64^). In the conditional models, difficult awakening that is independent of chronotype was associated with later mid-sleep timing (β_xy_ Conditional = 0.24, SE = 0.12, *p* = 0.053) and later timing of the most active 10 hours of the day (β_xy_Conditional = 0.24, SE = 0.12, *p* = 0.047). In order to allow comparison of effect sizes between the chronotype and difficulty awakening MR analyses we scaled the difficulty awakening MR effects to the ratio of the chronotype and difficulty awakening effects on sleep midpoint timing, thus placing them on the same scale. These scaled effects are reported in Figure 4 and Supplementary Table 19 alongside the unscaled effects.

**Figure 4.**
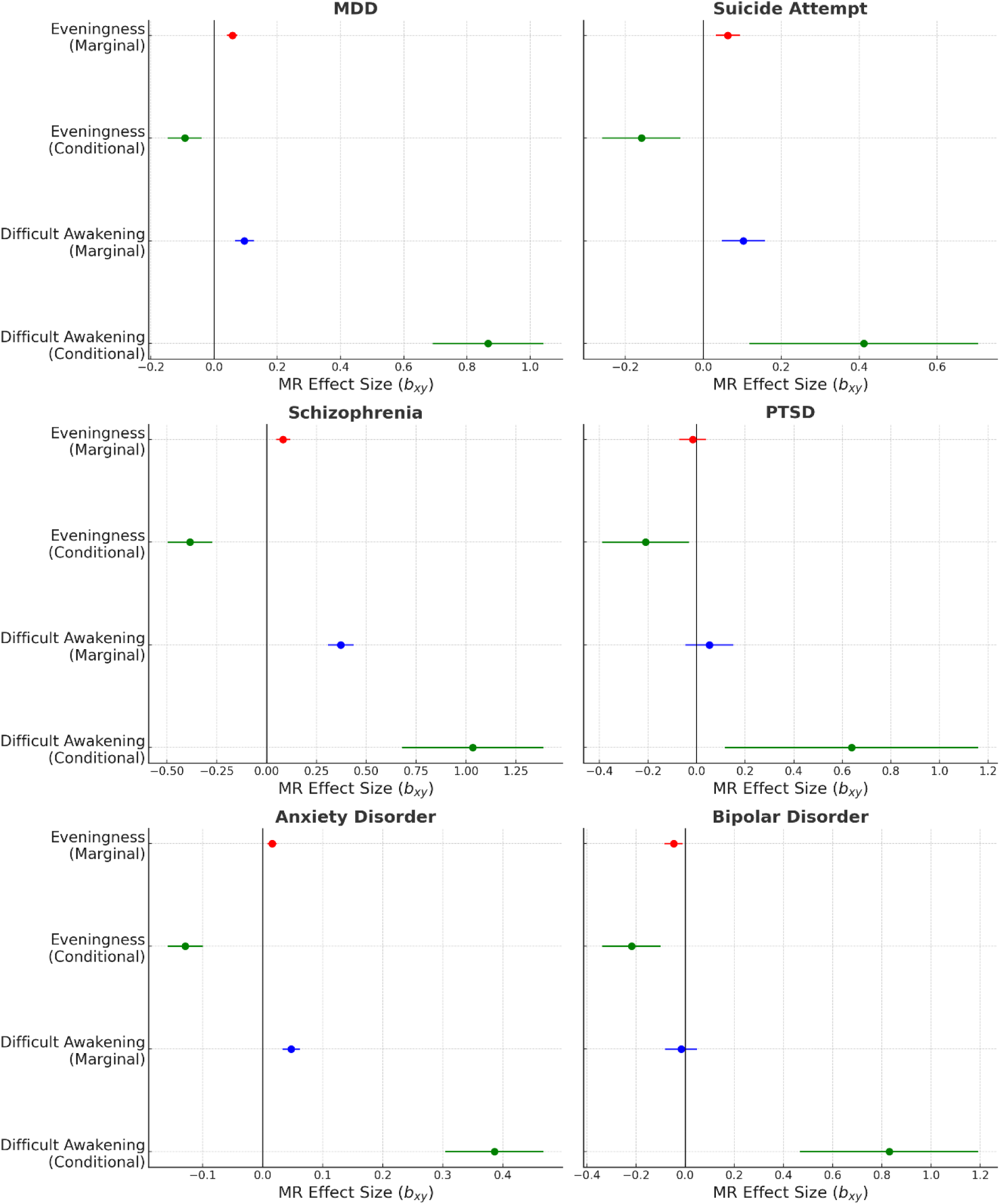
GSMR Mendelian Randomization effect size plots for the causal effects of difficult awakening an chronotype in marginal and conditional (mtCOJO adjusted) models on (A) major depressive disorder, (B) suicide attempt, (C) schizophrenia, (D) PTSD, (E) anxiety disorder and (F) bipolar disorder. Coefficients represent the MR effect (*b*_xy_) on the logit scale and 95% CIs for difficult awakening and late chronotype GWAS on each respectiv psychiatric outcome GWAS. Coefficients for the unadjusted MR are displayed in blue (difficult awakening) and re (evening chronotype) and coefficients for the conditional MR effect of difficult awakening and evening chronotype after mutual mtCOJO adjustment are displayed in green. Note: MR effect sizes and 95% CIs for difficult awakening are scaled to the MR effect of evening chronotype on sleep-midpoint to enable comparison of effect sizes. Unscale values are presented alongside scaled values in Supplementary Table 19.

The marginal and conditional Mendelian randomization of psychiatric traits and attempted suicide revealed a strikingly similar pattern of causal associations as that of the UK Biobank and OFTC epidemiological associations (Figure 4, Supplementary Table 19). In the marginal models, both genetically instrumented difficulty awakening and evening chronotype were causally associated with increased risk for MDD, suicide attempt, schizophrenia, and anxiety disorders (all *p* < 0.0005). Of note, in the marginal models, evening chronotype was causally associated with reduced risk for bipolar disorder (β_xy_ Marginal = –0.05, SE = 0.02, *p* = 0.01) while difficult awakening showed no association (*p* > 0.05). Further, neither difficulty awakening and evening chronotype were causally associated with PTSD (*p*_both_ > 0.05) in the marginal models. The conditional MR models revealed a striking separation in the conditional effects of evening chronotype and difficult awakening on psychiatric disorders and attempted suicide. Genetically instrumented evening chronotype that is independent of difficulty awakening was associated with *reduced* risk for MDD (*p* = 0.0008), attempted suicide (*p* = 0.002), schizophrenia (*p* = 1.84×10^−11^), PTSD (*p* = 0.02), and anxiety disorders (*p* = 9.34×10^−18^; Figure 4, Supplementary Table 19). Furthermore, in this conditional model the marginal association of evening chronotype with reduced risk for bipolar disorder was stronger (β_xy_Conditional = –0.22, SE = 0.06, *p* = 0.0003). In contrast, genetically instrumented difficult awakening that is independent of chronotype was causally associated with increased risk for MDD (*p* = 3.30×10^−22^), suicide attempt (*p* = 0.006), schizophrenia (*p* = 1.07×10^−8^), PTSD (*p* = 0.01), anxiety disorders (*p* = 2.30×10^−20^), and bipolar disorder (*p* = 7.19×10^−6^), with larger effect sizes as compared to the marginal models’ effect of difficult awakening on these disorders (Figure 4, Supplementary Table 19).

Lastly, we performed reverse MR for each of the outcome traits above to examine the magnitude and direction of effect of these traits on chronotype and difficulty awakening. Of the assessed traits, sleep midpoint timing, most active 10h timing, suicide attempt and PTSD GWAS had insufficient genome-wide significant loci (<10) for instrumental variable analysis and were therefore excluded. In reverse MR, MDD, schizophrenia, anxiety disorder and bipolar disorder were all associated with increased difficulty awakening in both marginal and conditional analyses. However, the strength of these associations was considerably weaker as compared to the forward analyses described above (Supplementary Table 20). For chronotype, while in the forward marginal and conditional models evening chronotype was associated with reduced risk for bipolar disorder, the opposite was observed in the reverse MR, such that bipolar disorder was associated with increased risk of evening chronotype (*p* < 0.0001; Supplementary Table 20). Furthermore, in the marginal reverse analysis MDD was not associated with increased risk for evening chronotype (*p* = 0.97), while the conditional association was of a similar strength and direction to the forward analysis (Supplementary Table 20).

## Discussion

The present study first aimed to examine whether sleep inertia drives the increased incidence of psychiatric disorders and suicide among evening chronotypes in longitudinal analyses of two large, independent cohorts (UK Biobank & Older Finnish Twin Cohort). Second, we aimed to complete a genome-wide association study of sleep inertia to examine its heritability and shared polygenicity with chronotype, other sleep phenotypes and psychiatric disorders. Finally, we aimed to use genetic instruments for sleep inertia and chronotype in marginal and conditional Mendelian randomization analyses to examine whether the genetically instrumented causal effect of evening chronotype on psychiatric disorders and suicide is explained by an underlying shared genetic architecture with sleep inertia.

We observed a moderate, positive correlation between measures of sleep inertia and eveningness in both the UK Biobank and Older Finnish Twin Cohort samples. This finding is at odds with the general conceptualization of chronotype and questionnaire measures of the construct which treat it as unitary and treats items related to sleep inertia and diurnal preference (morningness-eveningness) as interchangeable and belonging to a single common factor [10, 36] or in the case of the Munich Chronotype Questionnaire (MCTQ) do not include items relating to sleep inertia in their index of chronotype [28]. This finding is, however, consistent with numerous prior studies that have examined the factor structure of chronotype questionnaires and reliably identified sleep inertia and diurnal preference as distinct but non-orthogonal subordinate constructs of chronotype [26, 27, 61–64]. The finding that evening types tend to experience greater sleep inertia is expected given the known influence of circadian phase on sleep inertia, such that delayed circadian timing among evening types results in a shorter phase angle of awakening relative to the core body temperature minimum, i.e., misaligned wake in the biological night [22, 23].

An alternative explanation for this finding could be that evening types are at greater risk of poor-quality sleep due to sleep being mistimed, which could lead to increase sleep inertia due to higher homeostatic sleep pressure. Our findings do not support this explanation as we observed that the relationship between sleep inertia and evening chronotype could not be explained by self-reported sleep duration, insomnia symptoms or daytime sleepiness. Our analysis of objective sleep metrics from actigraphy mirrored this finding. We observed that later mid-sleep timing was associated with both greater sleep inertia and evening chronotype and that these associations were robust to adjustment for objectively measured sleep duration and sleep efficiency. Furthermore, while both evening chronotype and sleep inertia were associated with a reduced SRI in marginal analyses, in the conditional analyses only sleep inertia remained associated with a reduced SRI, indicating that the link between eveningness and irregular sleep wake patterns is driven by evening types with sleep inertia. This finding is meaningful as the SRI measures the day-to-day variability of sleep-wake patterns within an individual and is a marker of circadian rhythm misalignment with the sleep-wake cycle [39, 65]. As above, this pattern of associations was independent of objectively measured sleep duration and quality and therefore this finding is consistent with the interpretation that sleep inertia among evening types is in part an index of misaligned wake. Collectively, these findings suggest that sleep inertia is greater among evening types, those with later mid-sleep timing and those with sleep-wake rhythm disruption and that these associations are independent of subjective and objective measures of sleep quality, duration and daytime sleepiness.

Across ∼6 million person-years of observation in ∼400,000 participants from the UK Biobank, we observed that both evening chronotype and greater sleep inertia predicted an increased incidence of psychiatric disorders, including depression, schizophrenia, anxiety and bipolar disorders. These findings align with many prior studies linking evening chronotype with mood and psychotic disorders [1, 3, 5, 6, 8] and a smaller body of research that has identified sleep inertia as a common symptom across psychiatric disorder diagnoses that is strongly predictive of their prevalence and is associated with depressive symptoms in healthy individuals [66–69]. Here, we extend these findings, which have largely been drawn from small, cross-sectional studies, to a large prospective cohort design. We demonstrate that evening chronotype and greater sleep inertia at baseline in undiagnosed individuals predicts subsequent psychiatric disorder diagnosis across ∼16 years of follow-up. Importantly, these findings were robust to demographic, physical activity, sleep duration, sleep quality and daytime sleepiness covariates. This study also provides the first evidence that sleep inertia is longitudinally predictive of death by suicide across 38 years of follow-up in the Older Finnish Twin Cohort.

It is proposed that evening chronotypes are at higher risk for psychiatric disorders due to their increased risk for circadian misalignment, which is known to engender mood disruption [2, 16, 17, 70–74]. In accordance with this, misaligned wake that is too “early” for the circadian system, occurring in the biological night, is known to dramatically increase sleep inertia and evening chronotypes reliably experience greater sleep inertia compared to morning types, a phenomenon which is pronounced on work days when these individuals are challenged to wake early relative to the timing of their circadian system [22, 23, 26–28]. Given these findings, we sought to examine whether sleep inertia could be acting as a proxy for misaligned wake and therefore explain the association of evening chronotype with psychiatric disorders. Critically, we found that the effect of evening chronotype was mediated by sleep inertia for all outcomes, such that evening types without sleep inertia were at no greater risk as compared to morning types. In contrast, the effect of sleep inertia on incident psychiatric disorder risk remained unchanged when accounting for chronotype. This pattern of associations was robust to adjustment for lifestyle and demographic factors and in sensitivity analyses that accounted for sleep quality and daytime sleepiness as potential confounders and/or mediators of the association between evening chronotype and psychiatric disorders. This indicates that sleep inertia specifically explains the increased risk for developing psychiatric disorders among evening types rather than impaired sleep quality and distinguishes sleep inertia from daytime sleepiness as a biomarker of psychiatric disorder risk [75, 76]. These results are also in line with some small, cross-sectional studies which observed that sleep inertia acts as a mediator of the association between eveningness and depressed mood in non-clinical samples [61, 62, 77]. Importantly, we replicated these results in cross-sectional analyses of the same disorders in the UK Biobank and in longitudinal analyses of suicide and depressive symptoms in the Older Finnish Twin Cohort. These findings provide support for the interpretation that evening types are at greater risk for psychiatric disorders due to circadian misalignment, for which sleep inertia may be acting as a biomarker. Together, these results provide new evidence for sleep inertia as a driver of the associations between evening chronotype and the trans-diagnostic incidence of psychiatric disorders, including mood and psychotic disorders and suicide.

In analyses of twin and genome-wide association studies we established the heritability of sleep inertia, its enrichment in key genes and pathways, and its shared genetic architecture with chronotype, other sleep traits and psychiatric disorders. Twin analyses of the Older Finnish Twin Cohort established much stronger within-twin-pair correlations of sleep inertia for monozygotic twins as compared to dizygotic twins and twin modelling confirmed the trait to have a moderate broad-sense heritability of 40%, indicating both significant genetic and environmental contributors to the trait. This result is similar to those obtained in previous analyses of chronotype in the same cohort which we replicate here and extend by demonstrating that sleep inertia and chronotype share genetic and environmental causes [42]. Genome-wide common variant association analyses corroborated these findings, we observed significant narrow-sense heritability of sleep inertia of ∼8%, and we identified 98 independent genomic risk loci for the trait. Tissue and gene enrichment analyses demonstrated that sleep inertia loci were enriched in brain and pituitary tissues as well as known circadian rhythms genes, including *PER2, PER3, CRY1, CIART, FBXL3*; genes involved in ancillary pathways such as G protein-coupled receptor signaling (*NMUR2, RGS16, GPR61*); mitogen activated protein kinase signaling (*RASD1*, *MAPK12*); and synaptic organization (*NRXN1, SHANK2*). We also observed enrichment of difficulty awakening associations within the *RASA4* segmental duplication region, a modulator gene of MAPK signaling pathway which is critical for circadian photoentrainment, which has recently been implicated in evening chronotype and type 2 diabetes risk [78–80]. Bivariate Gaussian mixture modelling supported significant polygenic overlap of chronotype and sleep inertia, with most variants influencing one trait also influencing the other. However, there was also significant unshared polygenicity for both traits and simultaneously a significant degree of discordance among shared loci, with variants having diverging effects on the two traits, a finding which became important for subsequent conditional Mendelian randomization analyses. Finally, genetic correlations of sleep inertia and chronotype with sleep traits identified that while both show strong genetic correlations with the timing of mid-sleep and the most active 10h of the day, they have weak correlations with sleep duration, quality and daytime sleepiness. Of note, while late chronotype is not significantly genetically correlated with sleep duration or insomnia symptoms, sleep inertia shows a small genetic correlation with both longer sleep duration and increased insomnia symptoms, a pattern consistent with mis-timed sleep. With respect to psychiatric traits, genetic correlations with difficulty awakening and chronotype were positive, though with larger effect sizes for difficulty awakening in general.

Finally, we completed conditional GWAS of sleep inertia and chronotype which enabled us to examine both marginal and conditional causal associations of these traits with psychiatric disorders and suicide in Mendelian randomization analyses. In accordance with the bivariate mixture modelling, there was significant pleiotropy observed between chronotype and sleep inertia loci, such that conditioning one on the other in mtCOJO analyses dramatically reduced the number of genomic risk loci passing the genome-wide significance threshold. However, we did identify 24 genomic risk loci for sleep inertia that were independent of chronotype and 40 loci for chronotype that were independent of sleep inertia. Interestingly, eleven of these loci were shared between the conditional sleep inertia and chronotype GWAS due to discordant SNP effects. These included rs4757137 in the promoter region of the core clock gene *BMAL1* as well as rs6968865 within the *AHR* locus and rs16872808 within the *ZFPM2* locus, both of which have been linked to circadian functioning [81–83]. Critically, this conditional GWAS enabled us to pursue marginal and conditional two-sample MR analyses of sleep inertia, chronotype and psychiatric disorders by identifying genetic instruments for these traits that were not pleiotropic. First, we established that in both the marginal and conditional MR of chronotype on mid-sleep and most active 10h timing, evening chronotype was causally associated with later timing of these behaviors even when accounting for pleiotropy with sleep inertia. This is a critical finding as it indicates that our conditional MR of evening chronotype genetically proxies the delayed timing of sleep-wake behavior independent of sleep inertia.

The marginal and conditional MR analyses of psychiatric disorders remarkably mirrored the findings of the epidemiological analyses. In marginal analyses later chronotype is generally associated with increased risk for psychiatric disorders and attempted suicide, with bipolar disorder being a notable exception whereby later chronotype is associated with reduced risk even in the marginal analyses. In the conditional analyses we observed that shared genetic architecture with difficulty awakening drove these associations, such that genetically instrumented evening chronotype, which proxies later sleep-wake timing, is causally associated with reduced risk for each psychiatric disorder and attempted suicide when unconfounded with shared polygenicity with sleep inertia. This finding is notable as it dissociates later sleep-wake timing from psychiatric disorder risk absent sleep inertia. These findings challenge the idea that evening types with delayed sleep-wake timing are at increased risk for psychiatric disorders *per se*, instead suggesting that this effect is mediated by increased risk for circadian misaligned wake, for which sleep inertia is a known biomarker [7, 8, 22, 23, 84, 85]. Indeed, we observe that genetically proxied evening types with delayed sleep-wake timing that is uncoupled from sleep inertia are at a reduced risk for these disorders and attempted suicide. In contrast, sleep inertia is shown to be robustly causally associated with increased risk for each psychiatric disorder and attempted suicide. Importantly, reverse MR associations were considerably weaker than forward MR effects or absent altogether.

This study has several important limitations. First, while we replicate the observed pattern of associations in several studies using a diverse array of epidemiological and genetic methodologies, we should note that the measures of sleep inertia used in this study are subjective and objective (i.e., neurobehavioral) measures are known to differ in their time course and sensitivity to circadian phase/sleep loss [22, 24]. However, it has been shown that subjective measures of sleep inertia strongly predict their objective counterparts [76]. Secondly, it is important to acknowledge that the bulk of these results are drawn from the UK Biobank which is a cohort of middle and older adults and given the changes in circadian rhythms, sleep, and mood across the lifespan, our findings may conceivably differ in younger adults, adolescents and children. The findings from the OFTC, which mirror the UK Biobank and include a wider age range, attenuate this concern. Third, while our longitudinal epidemiological and Mendelian randomization analyses suggest an effect from sleep inertia to psychiatric disorders, it is important to recognize that some degree of reverse causality may still be present in our results due to the phenomenology of sleep inertia as a symptom of psychiatric disorders.

The present study is the first to establish that sleep inertia drives the increased incidence of psychiatric disorders and suicide among evening chronotypes in longitudinal analyses of two large, independent cohorts. We completed a genome-wide association study of sleep inertia, established its heritability and shared polygenicity with chronotype, other sleep phenotypes and psychiatric disorders. Finally, using genetic instruments for sleep inertia and chronotype in marginal and conditional Mendelian randomization analyses we corroborated our epidemiological findings. We observed that the causal association of evening chronotype with risk for psychiatric disorders and suicide is driven by shared genetic architecture with sleep inertia, such that evening chronotype that is independent of sleep inertia is causally associated with delayed sleep-wake timing but lower risk of psychiatric disorders. These findings challenge the notion that evening chronotype is a risk factor for psychiatric disorders *per se*, instead providing support for the understanding that evening types are at greater risk for psychiatric disorders due to circadian misalignment with an early-oriented society, for which sleep inertia may be acting as a biomarker.

## Data availability statement

The data used in this study are available in the UK Biobank resource subject to project approval by the UK Biobank Access Management Team. The authors of the present study are approved for access under application 6818. The OFTC data is not publicly available due to the restrictions of informed consent. However, the OFTC data is available through the Institute for Molecular Medicine Finland (FIMM) Data Access Committee (DAC) for authorized researchers who have IRB/ethics approval and an institutionally approved study plan. To ensure the protection of privacy and compliance with national data protection legislation, a data use/transfer agreement is needed, the content and specific clauses of which will depend on the nature of the requested data.

## Acknowledgements.

This research has been conducted using the UK Biobank Resource under application 6818. We would like to thank the participants and researchers from the UK Biobank who contributed or collected data. This work is funded by the following grant: R01HG012810 (JML).

