## Supplementary Methods, Figures and Tables 1-7 for "Sleep inertia drives the association of evening chronotype with psychiatric disorders: epidemiological and genetic evidence"

**Supplementary Table 1.** Participant characteristics across difficulty awakening and chronotype categories in the UK Biobank.

|  | **Easy Awakening (N=407,125)** | **Difficult Awakening (N=89,695)** | **Early  Chronotype (N=277,737)** | **Late  Chronotype (N=166,437)** |
| --- | --- | --- | --- | --- |
| **Age (years)** |  |  |  |  |
| Mean (SD) | 57.1 (7.97) | 53.9 (8.12) | 57.1 (7.92) | 55.7 (8.29) |
| Median [Q1, Q3] | 59.0 [51.0, 64.0] | 54.0 [47.0, 61.0] | 58.0 [51.0, 64.0] | 57.0 [49.0, 63.0] |
| **Sex** |  |  |  |  |
| Female | 212,661 (52.2%) | 57,796 (64.4%) | 156,416 (56.3%) | 91,152 (54.8%) |
| Male | 194,464 (47.8%) | 31,899 (35.6%) | 121,321 (43.7%) | 75,285 (45.2%) |
| **Body Mass Index** |  |  |  |  |
| Mean (SD) | 27.4 (4.66) | 27.7 (5.33) | 27.3 (4.73) | 27.6 (4.90) |
| Median [Q1, Q3] | 26.7 [24.2, 29.8] | 26.8 [24.0, 30.3] | 26.7 [24.1, 29.8] | 26.9 [24.2, 30.0] |
| **Employed** |  |  |  |  |
| No | 177,782 (43.7%) | 34,610 (38.6%) | 120,781 (43.5%) | 68,057 (40.9%) |
| Yes | 229,343 (56.3%) | 55,085 (61.4%) | 156,956 (56.5%) | 98,380 (59.1%) |
| **Townsend Deprivation** |  |  |  |  |
| Mean (SD) | -1.41 (3.03) | -0.838 (3.28) | -1.41 (3.03) | -1.18 (3.14) |
| Median [Q1, Q3] | -2.23 [-3.69, 0.335] | -1.66 [-3.41, 1.33] | -2.23 [-3.68, 0.347] | -2.01 [-3.58, 0.752] |
| **Smoker** |  |  |  |  |
| No | 263,864 (64.8%) | 61,375 (68.4%) | 182,669 (65.8%) | 107,346 (64.5%) |
| Yes | 143,261 (35.2%) | 28,320 (31.6%) | 95,068 (34.2%) | 59,091 (35.5%) |
| **Self-reported Insomnia** |  |  |  |  |
| Never/rarely | 103,569 (25.4%) | 16,115 (18.0%) | 66,167 (23.8%) | 40,248 (24.2%) |
| Sometimes | 198,341 (48.7%) | 38,327 (42.7%) | 133,470 (48.1%) | 78,238 (47.0%) |
| Usually | 104,922 (25.8%) | 35,170 (39.2%) | 77,965 (28.1%) | 47,844 (28.7%) |
| Missing | 293 (0.1%) | 83 (0.1%) | 135 (0.0%) | 107 (0.1%) |
| **Self-reported Sleep Duration (hours)** |  |  |  |  |
| Mean (SD) | 7.16 (1.05) | 7.17 (1.30) | 7.14 (1.07) | 7.19 (1.12) |
| Median [Q1, Q3] | 7.00 [7.00, 8.00] | 7.00 [6.00, 8.00] | 7.00 [7.00, 8.00] | 7.00 [7.00, 8.00] |
| Missing | 4643 (1.1%) | 2981 (3.3%) | 3348 (1.2%) | 2893 (1.7%) |
| **Physical Activity (MET Minutes)** |  |  |  |  |
| Mean (SD) | 2,940 (3540) | 2,280 (3080) | 2,970 (3550) | 2,590 (3270) |
| Median [Q1, Q3] | 1,770 [810, 3590] | 1,290 [558, 2710] | 1,790 [819, 3630] | 1,510 [693, 3110] |
| Missing | 31,101 (7.6%) | 10,569 (11.8%) | 20,062 (7.2%) | 14,435 (8.7%) |

**Supplementary Table 2. Participant characteristics across difficulty awakening and chronotype categories in the Older Finnish Twin Cohort.**

|  | **Easy Awakening (N=14,556)** | **Difficult Awakening (N=9,252)** | **Early  Chronotype (N=13,633)** | **Late  Chronotype (N=10,221)** |
| --- | --- | --- | --- | --- |
| **Age (years)** |  |  |  |  |
| Mean (SD) | 41.5 (13.5) | 38.8 (12.6) | 42.9 (13.9) | 37.5 (12.1) |
| **Sex** |  |  |  |  |
| Female | 7,318 (50.3%) | 5,074 (54.8%) | 7,082 (51.9%) | 5,365 (52.5%) |
| Male | 7,238 (49.7%) | 4,178 (45.2%) | 6,551 (48.1%) | 4,856 (47.5%) |
| **Body Mass Index** |  |  |  |  |
| Mean (SD) | 24.0 (3.4) | 23.5 (3.5) | 24.1 (3.5) | 23.4 (3.4) |
| **Employed** |  |  |  |  |
| Yes | 10,504 (72.2%) | 6,613 (71.5%) | 9,560 (70.1%) | 7,466 (73.0%) |
| No | 3,975 (27.3%) | 2,603 (28.1%) | 3,998 (29.3%) | 2,709 (26.5%) |
| Missing | 77 (0.5%) | 36 (0.4%) | 75 (0.6%) | 46 (0.5%) |
| **Smoker** |  |  |  |  |
| Yes | 4,144 (28.4%) | 3,339 (36.1%) | 3,704 (27.2%) | 3,746 (36.7%) |
| No | 10,085 (69.3%) | 5,694 (61.5%) | 9,596 (70.4%) | 6,250 (61.1%) |
| Missing | 327 (2.2%) | 219 (2.4%) | 333 (2.4%) | 225 (2.2%) |
| **Self-reported Sleep Duration** |  |  |  |  |
| Short (<7h) | 2,179 (15.0%) | 1,334 (14.4%) | 1,948 (14.3%) | 1,560 (15.3%) |
| Medium (7-8h) | 9,839 (67.6%) | 6,078 (65.7%) | 9,120 (66.9%) | 6,808 (66.6%) |
| Long (≥8.5h) | 2,459 (16.9%) | 1,802 (19.5%) | 2,480 (18.2%) | 1,813 (17.7%) |
| Missing | 79 (0.5%) | 38 (0.4%) | 85 (0.6%) | 40 (0.4%) |
| **Physical Activity  (MET Quintiles)** |  |  |  |  |
| Q1 | 2,325 (16.0%) | 1,525 (16.5%) | 2,239 (16.4%) | 1,637 (16.0%) |
| Q2 | 2,899 (19.9%) | 1,993 (21.5%) | 2,672 (19.6%) | 2,214 (21.7%) |
| Q3 | 2,940 (20.2%) | 2,022 (21.9%) | 2,803 (20.6%) | 2,162 (21.2%) |
| Q4 | 2,993 (20.6%) | 1,858 (20.1%) | 2,813 (20.6%) | 2,046 (20.0%) |
| Q5 | 3,108 (21.4%) | 1,729 (18.7%) | 2,809 (20.6%) | 2,021 (19.8%) |
| Missing | 291 (2.0%) | 125 (1.4%) | 297 (2.2%) | 141 (1.4%) |

**Supplementary Table 3. Cross-sectional associations of difficulty awakening and late chronotype with sleep and light exposure parameters in marginal and conditional models in the actigraphy subset (*n* = 86,772) of the UK Biobank**

|  | **Model 1**  **aBeta (95% CI)** | | | | **Model 2**  **aBeta (95% CI)^†^** | | | |
| --- | --- | --- | --- | --- | --- | --- | --- | --- |
|  | **Marginal Effects** | | **Conditional Effects** | | **Marginal Effects** | | **Conditional Effects** | |
|  | **Evening Chronotype** | **Difficult Awakening** | **Evening Chronotype** | **Difficult Awakening** | **Evening Chronotype** | **Difficult Awakening** | **Evening Chronotype** | **Difficult Awakening** |
| **Mid-Sleep Timing** | **0.50 (0.48 – 0.51)** | **0.33 (0.31 – 0.35)** | **0.46 (0.44 – 0.47)** | **0.14 (0.12 – 0.16)** | **0.49 (0.48 – 0.51)** | **0.33 (0.31 – 0.34)** | **0.45 (0.44 – 0.47)** | **0.14 (0.12 – 0.16)** |
| **Sleep Regularity Index** | **-0.06 (-0.08 – -0.04)** | **-0.10 (-0.12 – -0.08)** | **-0.03 (-0.05 – -0.01)** | **-0.08  (-0.11 – -0.06)** | **-0.03 (-0.04 – -0.01)** | **-0.06 (-0.08 – -0.04)** | -0.01 (-0.03 – 0.01) | **-0.05 (-0.07 – -0.03)** |
| **Sleep Efficiency** | **-0.11 (-0.13 – -0.09)** | **-0.13 (-0.15 – -0.11)** | **-0.09 (-0.10 – -0.07)** | **-0.09 (-0.11 – -0.07)** | **-0.10 (-0.12 – -0.09)** | **-0.12 (-0.14 – -0.10)** | **-0.08 (-0.10 –-0.06)** | **-0.08 (-0.10 – -0.06)** |
| **Sleep Duration** | **-0.04 (-0.06 – -0.03)** | **0.02 (0.003 – 0.04)** | **-0.06 (-0.07 – -0.04)** | **0.04 (0.02 – 0.06)** | **-0.06 (-0.08 – -0.05)** | **-0.0001 (-0.00 – -0.02)** | **-0.07 (-0.09 – -0.06)** | **0.03 (0.004– 0.05)** |
| **Daytime Light Exposure** | **-0.08 (-0.09 – -0.06)** | **-0.08 (-0.09 – -0.06)** | **-0.06 (-0.08 – -0.05)** | **-0.05 (-0.07 – -0.03)** | **-0.05 (-0.06 – -0.04)** | **-0.05 (-0.06 – -0.03)** | **-0.04 (-0.06 – -0.03)** | **-0.03 (-0.04– -0.01)** |
| **Night-time Light Exposure** | **0.12 (0.11 – 0.14)** | **0.08 (0.06 – 0.09)** | **0.11 (0.10 – 0.13)** | **0.03 (0.01 – 0.05)** | **0.12 (0.10 – 0.13)** | **0.07 (0.06 – 0.09)** | **0.11 (0.09 – 0.13)** | **0.03 (0.01 – 0.05)** |

Note: linear regression was used for all outcomes and standardized beta coefficients are presented with their 95% confidence intervals for the marginal and conditional effects of both evening chronotype and difficulty awakening relative to the morning chronotype and easy awakening referents, respectively. In marginal models, difficulty awakening and chronotype are included separately, whereas in conditional models difficulty awakening and chronotype are included together. Model 1 is adjusted for age, sex, and season. †Model 2 is additionally adjusted for employment and physical activity. Bolded coefficients indicate significance at the .05 level.

**Supplementary Table 4. Cox model longitudinal associations of difficulty awakening and evening chronotype with incident psychiatric disorders in marginal and conditional models in the UK Biobank**

|  | **Longitudinal Model 1**  **aHR (95% CI)** | | | | **Longitudinal Model 2**  **aHR (95% CI)^†^** | | | |
| --- | --- | --- | --- | --- | --- | --- | --- | --- |
|  | **Marginal Effects** | | **Conditional Effects** | | **Marginal Effects** | | **Conditional Effects** | |
|  | **Evening Chronotype** | **Difficult Awakening** | **Evening Chronotype** | **Difficult Awakening** | **Evening Chronotype** | **Difficult Awakening** | **Evening Chronotype** | **Difficult Awakening** |
| **Major depressive disorder** | **1.19 (1.16 – 1.23)** | **1.88 (1.83 – 1.94)** | 0.98 (0.95 – 1.01) | **1.86  (1.80 – 1.93)** | **1.19 (1.16 – 1.23)** | **1.82 (1.76 – 1.88)** | 0.99 (0.96 – 1.03) | **1.81 (1.74– 1.88)** |
| **Schizophrenia** | **1.40 (1.16 – 1.68)** | **3.00 (2.51 – 3.62)** | 0.95 (0.77 – 1.17) | **2.95 (2.28 – 3.83)** | **1.30 (1.06 – 1.58)** | **2.69 (2.20 – 3.28)** | 0.93 (0.75 – 1.16) | **2.68 (2.12 – 3.38)** |
| **Generalized anxiety disorder** | **1.21 (1.18 – 1.24)** | **1.80 (1.74 – 1.85)** | 1.00 (0.97 – 1.03) | **1.77 (1.71 – 1.83)** | **1.20 (1.17 – 1.24)** | **1.77 (1.71 – 1.82)** | 1.01 (0.98 – 1.04) | **1.73 (1.67 – 1.79)** |
| **Bipolar disorder** | **1.38 (1.20 – 1.59)** | **2.05 (1.77 – 2.38)** | 1.11 (0.95 – 1.29) | **1.95 (1.65 – 2.31)** | **1.37 (1.18 – 1.59)** | **1.91 (1.63 – 2.23)** | 1.13 (0.96 – 1.33) | **1.84 (1.53 – 2.20)** |

Note: Cox proportional hazard models were used for all outcomes and adjusted hazard ratios (aHRs) are presented with their 95% confidence intervals for the marginal and conditional effects of both evening chronotype and difficulty awakening relative to the morning chronotype and easy awakening referents, respectively. In marginal models, difficulty awakening and chronotype are included separately, whereas in conditional models difficulty awakening and chronotype are included together. Model 1 is adjusted for age, sex, and season. †Model 2 is additionally adjusted for employment and physical activity. Bolded coefficients indicate significance at the .05 level.

**Supplementary Table 5. Sleep duration and daytime sleepiness sensitivity analysis of the longitudinal association of difficulty awakening and evening chronotype with incident psychiatric disorders in the UK Biobank**

|  | **Longitudinal Model 3**  **aHR (95% CI)** | | | |
| --- | --- | --- | --- | --- |
|  | **Marginal Effects** | | **Conditional Effects** | |
|  | **Evening Chronotype** | **Difficult Awakening** | **Evening Chronotype** | **Difficult Awakening** |
| **Major depressive disorder** | **1.18 (1.14 – 1.22)** | **1.70 (1.63 – 1.76)** | 1.00 (0.97 – 1.04) | **1.68 (1.61 – 1.75)** |
| **Schizophrenia** | **1.26 (1.02 – 1.56)** | **2.29 (1.83 – 2.86)** | 0.89 (0.67 – 1.18) | **2.30 (1.78 – 2.97)** |
| **Generalized anxiety disorder** | **1.19 (1.15 – 1.22)** | **1.65 (1.60 – 1.71)** | 1.02 (0.99 – 1.05) | **1.62 (1.56 – 1.68)** |
| **Bipolar Disorder** | **1.35 (1.15 – 1.57)** | **1**.**65 (1**.**39 – 1**.**95)** | 1.18 (0.99 – 1.39) | **1**.**54 (1**.**27 – 1**.**87)** |

Note: Cox proportional hazard models were used for all outcomes and adjusted hazard ratios (aHRs) are presented with their 95% confidence intervals for the marginal and conditional effects of both evening chronotype and difficulty awakening relative to the morning chronotype and easy awakening referents, respectively. In marginal models, difficulty awakening and chronotype are included separately, whereas in conditional models difficulty awakening and chronotype are included together. Model 3 is adjusted for age, sex, season, employment, physical activity, sleep duration, sleep duration-squared and daytime sleepiness. Bolded coefficients indicate significance at the .05 level.

**Supplementary Table 6.** **Cross-sectional associations of difficulty awakening and late chronotype with psychiatric disorders in marginal and conditional models in the UK Biobank**

|  | **Cross-Sectional Model 1**  **aOR (95% CI)** | | | | **Cross-Sectional Model 2**  **aOR (95% CI)^†^** | | | |
| --- | --- | --- | --- | --- | --- | --- | --- | --- |
|  | **Marginal Effects** | | **Conditional Effects** | | **Marginal Effects** | | **Conditional Effects** | |
|  | **Evening Chronotype** | **Difficult Awakening** | **Evening Chronotype** | **Difficult Awakening** | **Evening Chronotype** | **Difficult Awakening** | **Evening Chronotype** | **Difficult Awakening** |
| **Major depressive disorder** | **1.47 (1.44 – 1.51)** | **3**.**62 (3**.**51 – 3**.**73)** | 1.01 (0.99 – 1.04) | **3.43 (3.32 – 3.55)** | **1.42 (1.39 – 1.46)** | **2.99 (2.90 – 3**.**10)** | **1.05 (1.02 – 1.08)** | **2.84 (2.75 – 2.95)** |
| **Self-harm** | **1.32 (1.26 – 1.39)** | **2.11 (1.99 – 2.22)** | 1.03 (0.97 – 1.09) | **2.05 (1.92 – 2.18)** | **1.22 (1.16 – 1.29)** | **1.65 (1.56 – 1.75)** | 1.02 (0.97 – 1.09) | **1.63 (1.52 – 1.73)** |
| **Generalized anxiety disorder** | **1.32 (1.26 – 1.38)** | **2.84 (2.71 – 2.98)** | **0**.**91 (0**.**86 – 0**.**96)** | **2.89 (2.74 – 3.06)** | **1.26 (1.20 – 1.32)** | **2.39 (2.27 – 2.51)** | **0**.**92 (0**.**88 – 0**.**97)** | **2.44 (2.29 – 2.58)** |
| **PTSD** | **1.36 (1.30 – 1.42)** | **2.69 (2.58 – 2.81)** | **0.95 (0.90 – 0.99)** | **2.69 (2.55 – 2.83)** | **1.28 (1.22 – 1.34)** | **2.30 (2.19 – 2.41)** | **0.95 (0.90 – 0.99)** | **2.32 (2.19 – 2.45)** |
| **Bipolar disorder** | **1.44 (1.34 – 1.55)** | **2.42 (2.24 – 2.61)** | 1.08 (0.99 – 1.18) | **2.30 (2.10 – 2.51)** | **1.35 (1.25 – 1.46)** | **2.16 (1.99 – 2.34)** | 1.06 (0.97 – 1.15) | **2.07 (1.89 – 2.28)** |
| **Psychotic experiences** | **1.23 (1.17 – 1.29)** | **1**.**74 (1**.**65 – 1**.**83)** | 1.03 (0.98 – 1.09) | **1**.**70 (1**.**59 – 1**.**80)** | **1.20 (1.14 – 1.26)** | **1.60 (1.51 – 1.69)** | 1.04 (0.98 – 1.09) | **1**.**56 (1**.**46 – 1**.**66)** |

Note: logistic regression was used for all outcomes and odds ratios (ORs) are presented with their 95% confidence intervals for the marginal and conditional effects of both evening chronotype and difficulty awakening relative to the morning chronotype and easy awakening referents, respectively. In marginal models, difficulty awakening and chronotype are included separately, whereas in conditional models difficulty awakening and chronotype are included together. Model 1 is adjusted for age, sex, and season. †Model 2 is additionally adjusted for employment and physical activity. Bolded coefficients indicate significance at the .05 level.

**Supplementary Table 7. Associations of difficulty awakening and late chronotype with psychiatric disorders in marginal and conditional models within the actigraphy sub-sample (*n* = 86,772) additionally adjusting for objectively measured sleep traits in the UK Biobank**

|  | **Cross-Sectional Model 3**  **aOR (95% CI)** | | | |
| --- | --- | --- | --- | --- |
|  | **Marginal Effects** | | **Conditional Effects** | |
|  | **Evening Chronotype** | **Difficult Awakening** | **Evening Chronotype** | **Difficult Awakening** |
| **Major depressive disorder** | **1.29 (1.23 – 1.34)** | **2.49 (2.36 – 2.62)** | 1.01 (0.96 – 1.05) | **2.41 (2.28 – 2.57)** |
| **Self-harm** | **1.18 (1.08 – 1.28)** | **1.65 (1.51 – 1.80)** | 0.99 (0.90 – 1.09) | **1.65 (1.49 – 1.83)** |
| **Generalized anxiety disorder** | **1.15 (1.07 – 1.25)** | **2.84 (2.71 – 2.98)** | **0**.**86 (0**.**79 – 0**.**94)** | **2.34 (2.15 – 2.60)** |
| **PTSD** | **1.21 (1.12 – 1.30)** | **2.18 (2.02 – 2.35)** | **0.91 (0.83 – 0.98)** | **2.26 (2.06 – 2.47)** |
| **Bipolar disorder** | **1.31 (1.15 – 1.49)** | **1.92 (1.67 – 2.20)** | 1.08 (0.94 – 1.23) | **1.86 (1.59 – 2.18)** |
| **Psychotic experiences** | **1.19 (1.10 – 1.29)** | **1**.**62 (1**.**49 – 1**.**77)** | 1.03 (0.94 – 1.12) | **1**.**60 (1**.**45 – 1**.**77)** |

Note: logistic regression was used for all outcomes and adjusted odds ratios (aORs) are presented with their 95% confidence intervals for the marginal and conditional effects of both evening chronotype and difficulty awakening relative to the morning chronotype and easy awakening referents, respectively. In marginal models, difficulty awakening and chronotype are included separately, whereas in conditional models difficulty awakening and chronotype are included together. Model 3 is adjusted for age, sex, season, employment as well as objectively measured physical activity, sleep duration, sleep efficiency and sleep midpoint. Bolded coefficients indicate significance at the .05 level.

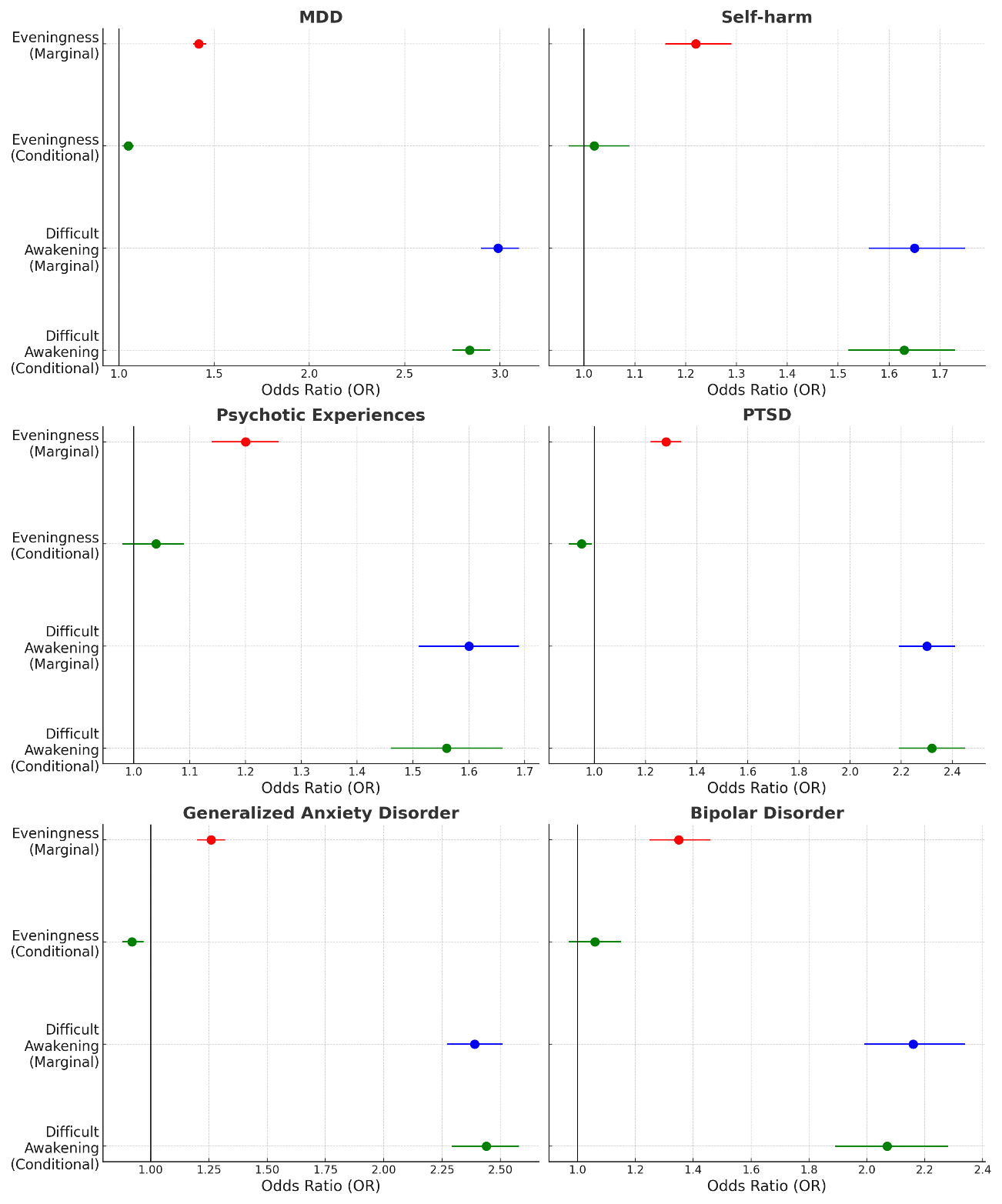

**Supplementary Figure 1.** Coefficients plot for the marginal and conditional cross-sectional associations of difficult awakening and late chronotype with (A) major depressive disorder, (B) self-harm behavior, (C) psychotic experiences, (D) PTSD, (E) generalized anxiety disorder and (F) bipolar disorder. Coefficients represent the odds ratios ± the 95% CI for difficulty awakening and late chronotype relative to easy awakening and early chronotype referents, respectively. Model 2 is presented adjusted for age, sex, ethnicity, season, employment, and physical activity. Coefficients for the marginal effects are displayed in blue (difficult awakening) and red (late chronotype) and coefficients for the conditional effect of difficult awakening and late chronotype after mutual adjustment are displayed in green.

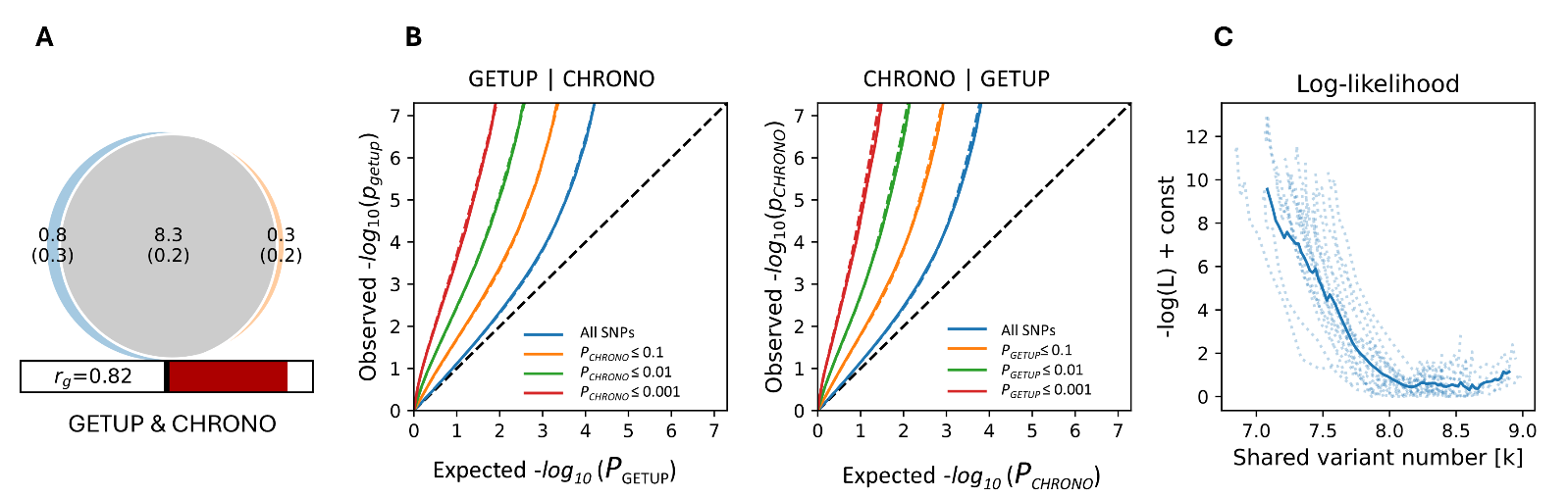

**Supplementary Figure 2.** MiXeR analysis results. (A) Venn diagram depicting the shared and unshared polygenicity of difficulty awakening and chronotype with values in thousands of variants (SD) and a bar graph of the genetic correlation. (B) Conditional Q-Q plots depicting the cross-trait SNP enrichment between difficulty awakening and chronotype indicating shared genetic architecture. (C) Negative log-likelihood plot indicating model convergence.

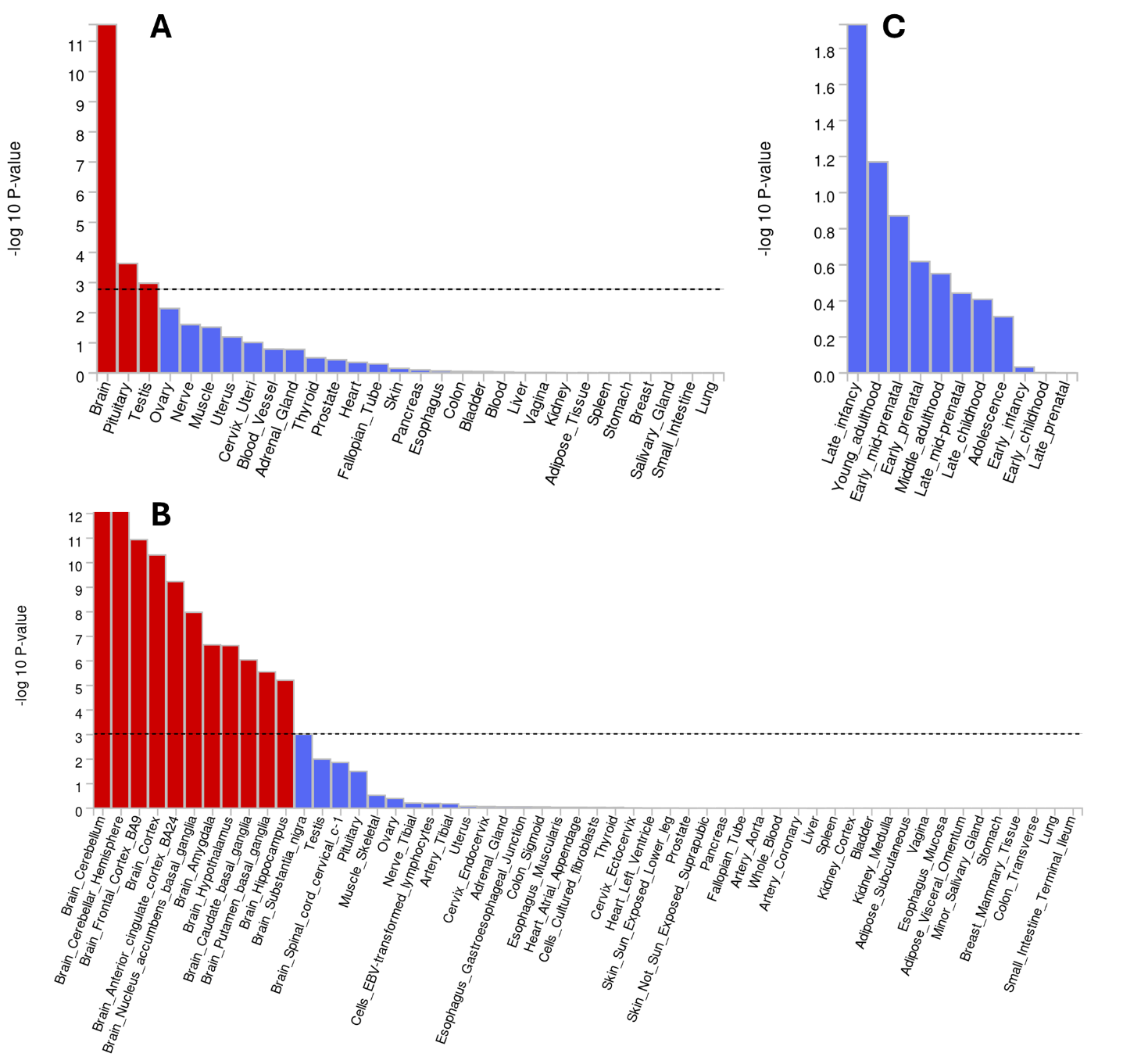

**Supplementary Figure 3.** MAGMA tissue enrichment of difficulty awakening loci among (A) superordinate and subordinate (B) human tissues and (C) across 11 developmental stages of brain samples. Red highlighted bars indicate tissues Bonferroni-significantly enriched for difficulty awakening loci.

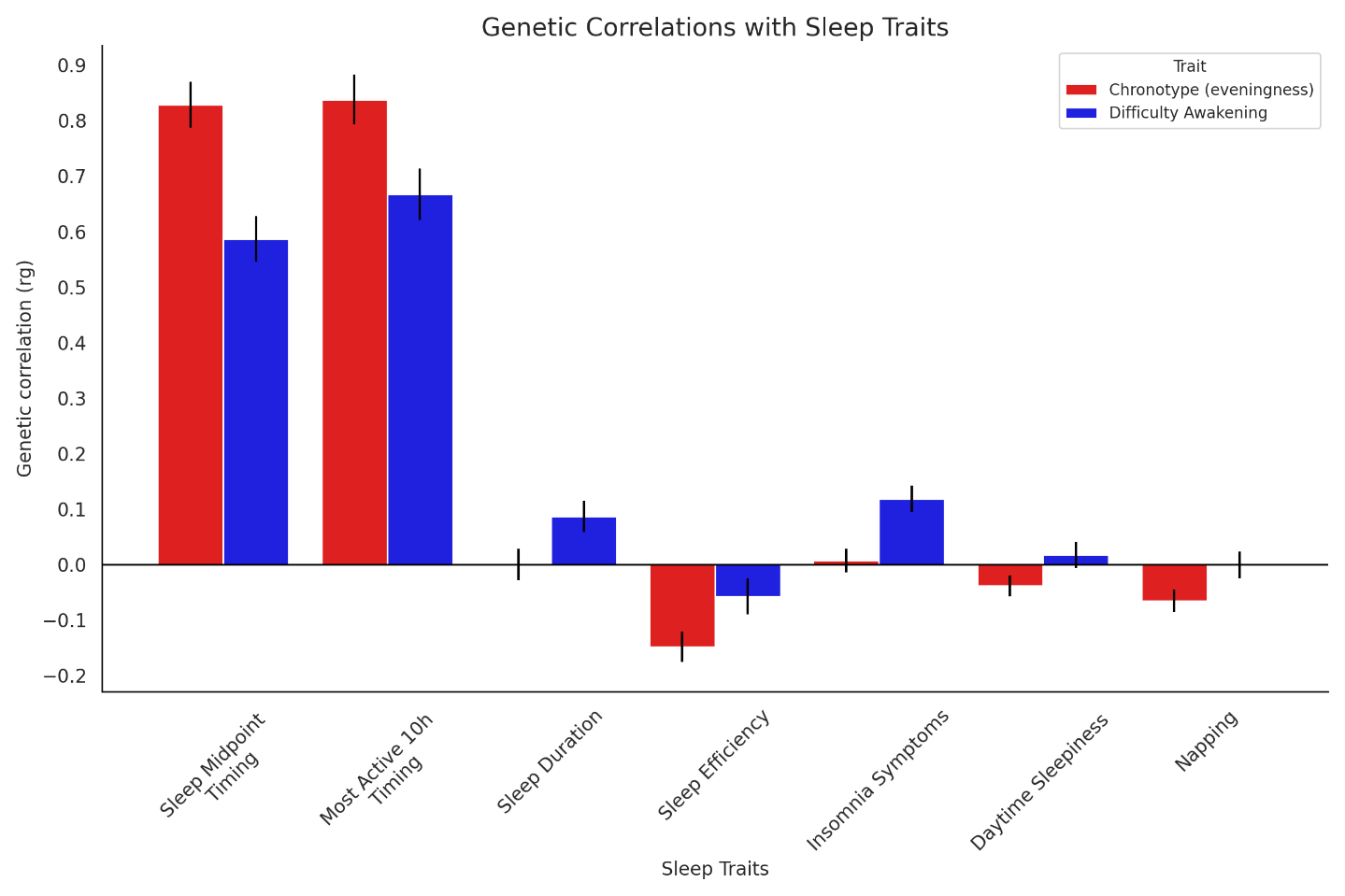

**Supplementary Figure 4.** LDSC genetic correlations (*r*_g_) of difficulty awakening and chronotype with sleep traits. Error bars represent standard error of the mean.

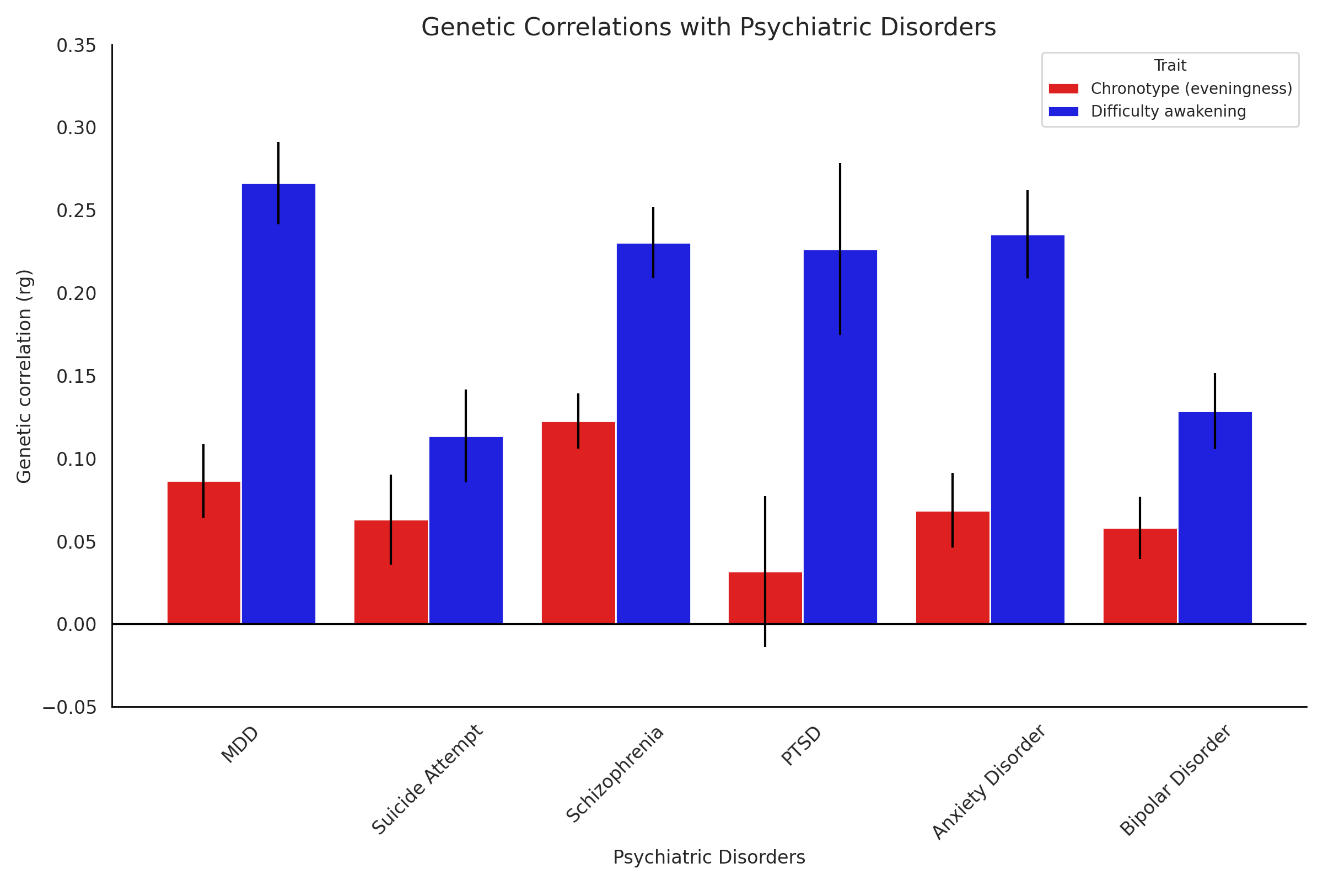

**Supplementary Figure 5.** LDSC genetic correlations (*r*_g_) of difficulty awakening and chronotype with psychiatric disorders and suicide attempt. Error bars represent standard error of the mean.

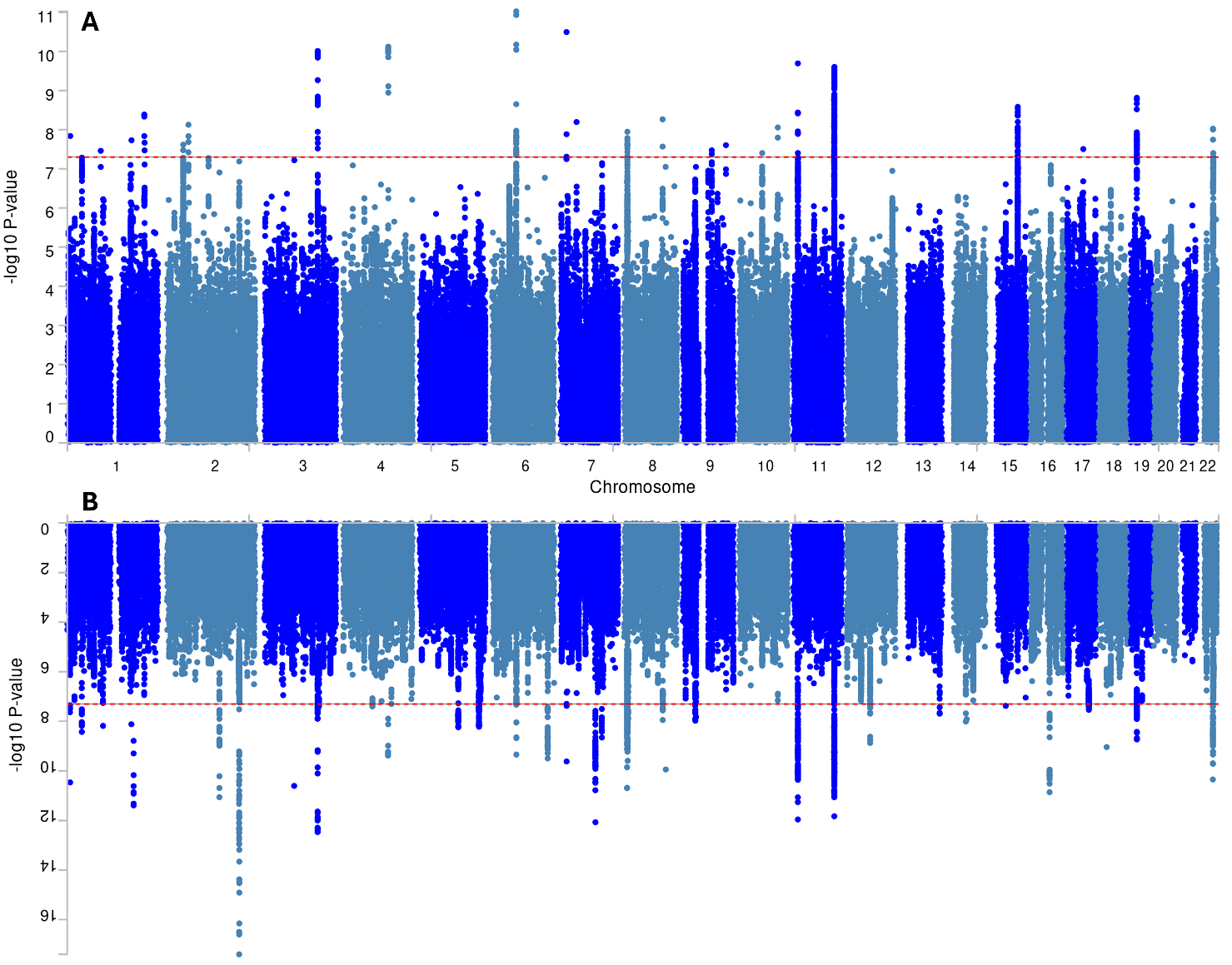

**Supplementary Figure 6.** Miami plot of the mtCOJO conditional GWAS of (A) difficulty awakening conditioned on chronotype and (B) chronotype conditioned on difficulty awakening.

**Supplementary Methods.**

**Sleep inertia and chronotype definitions in the UK Biobank and Older Finnish Twin Cohort**

In the UK Biobank cohort, sleep inertia was measured by self-reported difficulty awakening (data-field 1170) and chronotype was measured by self-reported diurnal preference (data-field 1180) as part of the baseline questionnaire between 2006 to 2010. Both items were drawn from the Horne-Ostberg Morningness-Eveningness Questionnaire (Horne & Ostberg, 1976). For difficulty awakening, participants were prompted to answer the question “On an average day, how easy do you find getting up in the morning?” with one of six possible answers: “Not at all easy”, “Not very easy”, “Fairly easy”, “Very easy”, “Do not know” or “Prefer not to answer”. For chronotype, participants were prompted to answer the question “Do you consider yourself to be?” with one of six possible answers: “Definitely a ‘morning’ person”, “More a ‘morning’ than ‘evening’ person”, “More an ‘evening’ than a ‘morning’ person”, “Definitely an ‘evening’ person”, “Do not know” or “Prefer not to answer”. Responses of “Do not know” and “Prefer not to answer” were coded as missing for both items. The UK Biobank difficulty awakening and chronotype items were binarized as follows for analysis: difficulty awakening (0 = “Very easy” or “Fairly easy” vs. 1 = “Not very easy” or “Not at all easy”) and chronotype (0 = “Definitely a morning person” or “More a ‘morning’ than ‘evening’ person”; 1 = “More an ‘evening’ than a ‘morning” person” or “Definitely an evening person”).

The OFTC study measured sleep inertia with an item on difficulty awakening and chronotype with an item on diurnal preference, both taken from the Diurnal Type Scale in the 1981 questionnaire (Torsvall & Åkerstedt, 1980). For difficulty awakening, participants were asked “How long does it take to you to feel fully alert and functioning in the morning after a night’s sleep?” with one of four response options: “less than 10 minutes”, “10-19 minutes”, “20 to 39 minutes” or “more than 40 minutes”. For chronotype, participants were asked to “Try to assess to what extent you are a morning person or an evening person” with four response options: “I am clearly a morning person”, “I am to some extent a morning person”, “I am to some extent an evening person” or “I am clearly an evening person”. The OFTC difficulty awakening and chronotype items were binarized as follows for analysis: difficulty awakening (0 = “less than 10 minutes” vs. 1 = “10-19 minutes” or “20-39 minutes” or “more than 40 minutes”) and chronotype (0 = “I am clearly a morning person” or “I am to some extent a morning person”; 1 = “I am to some extent an evening person” or “I am clearly an evening person”).

**Genetic ancestry definition and principial components of ancestry**

European ancestry classification of UK Biobank participants for the GWAS analysis was undertaken using the Human Genome Diversity Project-1000 Genomes (HGDP-1KG) harmonized reference dataset (Koenig et al., 2024)_._ The HGDP-1KG is a high-quality dataset of 4,094 whole genomes from labelled diverse continental populations. Principal components analysis (PCA) was first performed on unrelated (KING kinship coefficient < 0∙125) individuals in the HGDP-1KG dataset after pruning variants (500kb window, *r*^2^ = 0.10) to extract the top 10 PCs of ancestry (Manichaikul et al., 2010; Patterson, Price, & Reich, 2006; Purcell et al., 2007). We then projected individuals from the UK Biobank onto the HGDP-1KG PC space and trained a random forest classifier given continental ancestry labels from the HGDP-1KG cohort to assign ancestry to UK Biobank individuals based on their top 10 PC scores using the Python package *sklearn* (Pedregosa et al., 2011). The minimum random forest probability for assignment to a particular ancestry group was 0.5 and we completed 20 iterations of this model. An individual was assigned to the European ancestry group and included in the genome-wide association study of difficulty awakening if 20/20 iterations assigned them to the European ancestry, otherwise individuals were excluded as non-European or admixed. Finally, PCA was then completed within European UKB individuals to extract the top 10 PCs of ancestry for inclusion in the GWAS as covariates to adjust for population stratification within the European sub-population.

**References for external GWAS of sleep-wake, psychiatric disorder and attempted suicide traits**

| **GWAS Trait** | **Reference** |
| --- | --- |
| Chronotype | (Jones, Lane, et al., 2019) |
| Mid-sleep timing | (Jones, van Hees, et al., 2019) |
| Most active 10h timing | (Jones, van Hees, et al., 2019) |
| Sleep duration | (Jones, van Hees, et al., 2019) |
| Sleep efficiency | (Jones, van Hees, et al., 2019) |
| Insomnia symptoms | (Lane et al., 2019) |
| Daytime sleepiness | (Wang et al., 2019) |
| Napping | (Dashti et al., 2021) |
| MDD | (Als et al., 2023) |
| Suicide Attempt | (Docherty et al., 2023) |
| Schizophrenia | (Trubetskoy et al., 2022) |
| PTSD | (Nievergelt et al., 2019) |
| Anxiety Disorder | (Friligkou et al., 2024) |
| Bipolar Disorder | (Mullins et al., 2021) |

**Accelerometry and sleep analysis**

The following methods are adapted from Burns et al. (2023). In 2013, 236,519 UK Biobank participants were invited to wear an accelerometer for 7 days as part of a physical activity and light monitoring study. Of these participants, 103,720 (43.9%) accepted, and returned the accelerometer to UK Biobank. Participants who accepted the invitation received a wrist-worn AX3 triaxial accelerometer (Axivity, Newcastle upon Tyne, UK) with in-built light sensor (APDS9007 silicon photodiode sensor; spectral sensitivity 𝜆 = 470-650nm) and were asked to wear the device on their dominant wrist for seven days under free-living conditions. At the end of the 7-day period, participants were instructed to return the accelerometer to UK Biobank using a prepaid envelope.

The R package GGIR (Doherty et al., 2017; van Hees et al., 2018) (v1.6-9) was used to assess analyse accelerometry data, assess data quality and provide summaries of device non-wear and sleep parameters.

1. The raw accelerometry data files for each individual were downloaded in Continuous Wave Accelerometer (cwa) format and converted to Waveform Audio File (wav) format using the open-source software OMConvert.
2. Periods of device non-wear were identified algorithmically by GGIR (van Hees et al., 2013) and excluded from analysis
3. Sleep periods were then determined by GGIR using a validated, heuristic algorithm(van Hees et al., 2018), briefly:
a) The median z-angle (perpendicular to the wrist) was calculated in 5 second epochs using the three perpendicular axes of motion in relation to the downward force of gravity
b) A 5-minute rolling median of the absolute differences in z-angle between the 5 second epochs was calculated
c) The 10th percentile of the rolling median across a day (from noon to noon) was calculated and multiplied by 15 to set a threshold for determining inactivity
d) Blocks of inactivity were those for which the rolling median z-angle difference (from step b) is less than the threshold (from step c) and which last for 30 or more minutes
e) Blocks of inactivity (from step d) less than 60 minutes apart were treated as the same inactivity block
f) Labelling the longest combined period of sustained inactivity (from step e) in each day (noon-noon) as the sleep period time windows (SPT window) and all other periods of sustained inactivity as diurnal (daytime) inactivity. Note: if the SPT ends after noon (indicating sleep offset after noon) the sleep analysis is repeated on a 6am-6pm window in order to detect daytime sleepers.
g) Participants without any valid days of sleep-wake data due to file corruption or consistent non-wear interfering with SPT window detection were excluded (*n* = 8,055)
h) Sleep duration was calculated as the duration of GGIR-determined sustained inactivity between sleep onset and offset times. Where, sustained inactivity was defined as less than 5 degrees of movement from the accelerometer z-axis across rolling 5-minute intervals (Windred et al., 2021).
i) Sleep efficiency was calculated as the proportion of sustained inactivity between sleep onset and offset, i.e. [total sustained inactivity between sleep onset and offset / (sleep offset time - sleep onset time)].
j) Sleep midpoint was calculated as the midpoint between sleep onset and offset times in hours after previous midnight.
k) Sleep regularity was assessed using the sleep regularity index (SRI), a metric that describes the stability of sleep-wake patterns from one day to the next (Phillips et al., 2017; Windred et al., 2021). The SRI calculates the average concordance in sleep–wake state of all epoch pairs separated by 24 hours. An SRI of 100 represents perfectly regular sleep–wake patterns, and zero represents random patterns.

**Light analysis**

Following GGIR analysis, participants light data was mapped to the GGIR output and analysed with custom R scripts. Participants lacking any valid days of sleep-wake data due to data file corruption or consistent non-wear identified by GGIR were also excluded from light analysis (as noted above, *n* = 8,055).

1. Participants’ light data were extracted from the cwa files.
2. Output current was then converted from the logarithmic scale to approximate lux according to the device manual (lux = 10^(device output current/341).
3. Output current of the light sensor was down-sampled from 100Hz to 1Hz and then averaged into 10-second epochs.
4. Epochs of device non-wear identified by GGIR were marked as missing data.
5. Participants with consistently dark (percentage of data below 10lux > 80%) or consistently bright (percentage of data above 6000lux >75%) were excluded (*n* = 7,651), indicating device malfunction or coverage.
6. Daily light profiles of participants were constructed by averaging epochs into forty-eight 30-min bins across the 24-hour period per study day and these bins were then averaged across the study days to generate an average 24-hour light exposure profile per-participant (i.e. each 30-minute bin contained the average light exposure at that time across the available study days).
7. Participants with less than two days’ worth of light data per 30-minute bin were excluded (*n* = 1,242) for a final sample of 86,772.

Light exposure predictors were defined by factor analysis of the forty-eight 30-minute light bins across the 24-hour period. Factor analysis supported the extraction of a two-factor structure (day, 7.30am-8.30pm; and night, 12.30am-6am; factor loadings ≥ 0.5, varimax rotation, cumulative proportion of variance explained = 0.56) based on the scree plot, the additional proportion of variance explained by each factor (>0.10), and the independence of resultant factors. Day and night light variables were calculated by averaging light values across the respective time bins. Internal consistency of both day (Cronbach’s α = 0.98) and night (α = 0.93) light variables was very good.

Reliability of day and night light measurements was assessed in a subset of the actigraphy sample (*n* = 2,988) that completed a series of four repeated actigraphy assessments. These repeated assessments occurred on average 3.23 years (SD = 0.65) after the main assessment and took place over the course of a year with three months between each measurement. A linear mixed-effects model with a crossed random effect structure including participant, assessment (one to four), and time of day (day and night) was used to estimate the intra-class correlation (ICC) of (log-transformed) light. Due to the large positive skew in both day and night light variables (skewness > 1 for both), both variables were converted into categorical predictors for analysis by dividing them into four equal-sized quartiles (*n*_Q1-Q4_ = 21,693) in ascending brightness. AX3 device output was calibrated to lux using the validation procedures and formula described in (Burns et al., 2023).

**UK Biobank cross-sectional psychiatric outcome variable definitions**

Psychiatric cross-sectional outcomes were defined based on participant responses to the UK Biobank Mental Health Questionnaire (MHQ) following guidelines established by Davis et al. (2020).

***Lifetime major depressive disorder***

Lifetime major depressive disorder cases were defined according to the Composite International Diagnostic Interview (CIDI) lifetime depression module based on the DSM definition of major depressive disorder as well as professional diagnosis of major depression(Kessler, Andrews, Mroczek, Ustun, & Wittchen, 1998).

*Cases*

- Responded ‘yes’ to either or both core symptom questions:
  - “Have you ever had a time in your life when you felt sad, blue, or depressed for two weeks or more in a row?? (data-field 20446)
  - “Have you ever had a time in your life lasting two weeks or more when you lost interest in most things like hobbies, work, or activities that usually give you pleasure?” (data-field 20441).
- AND responded “most of the day” or “all day long” to the question: “How much of the day did these feelings usually last?” (data-field 20436)
- AND responded “almost every day” or “every day” to the question: “Did you feel this way?” (data-field 20439)
- AND responded “somewhat” or “a lot” to the question: "Think about your roles at the time of this episode, including study / employment, childcare and housework, leisure pursuits. How much did these problems interfere with your life or activities?" (data-field 20440)
- AND endorsed ≥ 5 symptoms:
  - “Have you ever had a time in your life when you felt sad, blue, or depressed for two weeks or more in a row?” (data-field 20446)
  - “Have you ever had a time in your life lasting two weeks or more when you lost interest in most things like hobbies, work, or activities that usually give you pleasure?” (data-field 20441)
  - “Did you feel more tired out or low on energy than is usual for you?” (data-field 20449)
  - “Did you gain or lose weight without trying, or did you stay about the same weight?” (data-field 20536). Responding “gained”, “lost” or “both gained and lost” counted as endorsement.
  - “Did your sleep change?” (data-field 20532)
  - “Did you have a lot more trouble concentrating than usual?” (data-field 20435)
  - “People sometimes feel down on themselves, no good, worthless. Did you feel this way?” (data-field 20450)
  - “Did you think a lot about death - either your own, someone else's or death in general?” (data-field 20437)
- OR reported a professional diagnosis of major depression (data-field 20544 = 11)

*Controls*

- Responded “no” to both core symptom questions:
  - “Have you ever had a time in your life when you felt sad, blue, or depressed for two weeks or more in a row?” (data-field 20446)
  - “Have you ever had a time in your life lasting two weeks or more when you lost interest in most things like hobbies, work, or activities that usually give you pleasure?” (data-field 20441).
- Did not report a professional diagnosis of depression (data-field 20544)
- Had a PHQ-9 score of ≤ 5

***Lifetime generalized anxiety disorder***

Lifetime generalized anxiety disorder cases according to the CIDI lifetime GAD module based on the DSM-IV definition of GAD(Kessler et al., 1998).

*Cases*

- Responded yes to the question: “Have you ever had a period lasting one month or longer when most of the time you felt worried, tense, or anxious?” (data-field 20421)
- AND responded ≥ 6 months or “all my life/as long as I can remember” to the question: “What is the longest period of time that this kind of worrying has ever continued?” (data-field 20420)
- AND responded “yes” to the question: “Please think of the period in your life when you have felt worried, tense, anxious, or more worried than most people would in your situation. This could be in the past, or it could be continuing now. Did you worry most days?” (data-field 20538)
- AND responded
  - “more than most” to the question: “People differ a lot in how much they worry about things. Did you ever have a time when you worried a lot more than most people would in your situation?” (data-field 20425)
  - OR “stronger than most” to the question: “Please think of the period in your life when you have felt worried, tense, anxious, or more worried than most people would in your situation. This could be in the past, or it could be continuing now. During that period, was your worry stronger than in other people?” (data-field 20542)
- AND responded
  - “more than one thing” to the question: “Please think of the period in your life when you have felt worried, tense, anxious, or more worried than most people would in your situation. This could be in the past, or it could be continuing now. Did you usually worry about one particular thing, such as your job security or the failing health of a loved one, or more than one thing?” (data-field 20543)
  - OR “yes” to the question: “Please think of the period in your life when you have felt worried, tense, anxious, or more worried than most people would in your situation. This could be in the past, or it could be continuing now. Did you ever have different worries on your mind at the same time?” (data-field 20540)
- AND responded:
  - “yes” to the question: “Please think of the period in your life when you have felt worried, tense, anxious, or more worried than most people would in your situation. This could be in the past, or it could be continuing now. Did you find it difficult to stop worrying?” (data-field 20541)
  - OR “sometimes“ or “often” to the question: “Please think of the period in your life when you have felt worried, tense, anxious, or more worried than most people would in your situation. This could be in the past, or it could be continuing now. How often was your worry so strong that you couldn't put it out of your mind no matter how hard you tried?” (data-field 20539)
  - OR “sometimes” or “often” to the question: “Please think of the period in your life when you have felt worried, tense, anxious, or more worried than most people would in your situation. This could be in the past, or it could be continuing now. How often did you find it difficult to control your worry?” (data-field 20537)
- AND responded “somewhat” or “a lot” to the question: “Think about your roles at the time of this episode, including study / employment, childcare and housework, leisure pursuits. How much did these problems interfere with your life or activities?” (data-field 20418)
- AND endorsed three or more somatic symptoms by responding “yes” to the questions: “When you were worried or anxious, were you also:”
  - “Restless?” (data-field 20426)
  - “Keyed up or on edge?” (data-field 20423)
  - “Easily tired?” (data-field 20429)
  - “Having difficulty keeping on your mind what you were doing?” (data-field 20419)
  - “More irritable than usual?” (data-field 20422)
  - “Having tense, sore or aching muscles?” (data-field 20417)
  - “Often having trouble falling or staying asleep?” (data-field 20427)

*Controls*

- Did not meet the above criteria
- GAD-7 score of < 5

***Lifetime bipolar disorder (mania/hypomania)***

Lifetime bipolar disorder cases were defined according to DSM-IV guidelines(Carvalho et al., 2015; Cerimele, Chwastiak, Dodson, & Katon, 2014).

*Cases*

- Responded “yes” to either or both core symptom questions:
  - “Have you ever had a period of time when you were feeling so good, "high", "excited", or "hyper" that other people thought you were not your normal self or you were so "hyper" that you got into trouble?” (data-field 20501)
  - “Have you ever had a period of time when you were so irritable that you found yourself shouting at people or starting fights or arguments?” (data-field 20502)
- AND endorsed four or more features of bipolar disorder from:
  - “I was more active than usual” (data-field 20548(1))
  - “I was more talkative than usual” (data-field 20548(2))
  - “I needed less sleep than usual” (data-field 20548(3))
  - “I was more creative or had more ideas than usual” (data-field 20548(4))
  - “I was more restless than usual” (data-field 20548(5))
  - I was more confident than usual (data-field 20548(6))
  - “My thoughts were racing” (data-field 20548(7))
  - “I was easily distracted” (data-field 20548(8))
  - “Have you ever had a period of time when you were feeling so good, "high", "excited", or "hyper" that other people thought you were not your normal self or you were so "hyper" that you got into trouble?” (data-field 20501)
- AND responded “a week or more” when asked: “What is the longest time that these "high" or "irritable" periods have lasted?” (data-field 20492)
- AND responded “needed treatment or caused problems with work, relationships, finances, the low or other aspects of life” to the question: “How much of a problem have these "high" or "irritable" periods caused you?” (data-field 20493)
- OR reported a professional diagnosis of bipolar disorder (data-field 20544 = 10)

*Controls*

- Did not meet the above criteria
- Did not report a professional diagnosis of bipolar disorder (data-field 20544 = 10)

***Lifetime psychotic experiences***

Lifetime psychosis cases were defined using participant responses to the MHQ items which were adapted from the CIDI(Nuevo et al., 2010).

*Cases*

- Responded “yes” to any of the following questions:
  - “Did you ever believe that a strange force was trying to communicate directly with you by sending special signs or signals that you could understand but that no one else could understand (for example through the radio or television)?” (data-field 20474)
  - “Did you ever believe that there was an unjust plot going on to harm you or to have people follow you, and which your family and friends did not believe existed?” (data-field 20468)
  - “Did you ever see something that wasn't really there that other people could not see?” (data-field 20471)
  - “Did you ever hear things that other people said did not exist, like strange voices coming from inside your head talking to you or about you, or voices coming out of the air when there was no one around?” (data-field 20463)
- OR reported a professional diagnosis of schizophrenia (data-field 20544 = 2) or psychotic illness (data-field 20544 = 3)

*Controls*

- Did not endorse any of the four psychotic experiences
- Did not report a professional diagnosis of schizophrenia (data-field 20544 = 2) or psychotic illness (data-field 20544 = 3)

***Post-traumatic stress disorder***

PTSD cases and controls were defined using the PTSD-Checklist-6 (PCL-6) with a clinical case-control threshold of 14 that is validated against the CIDI(Lang & Stein, 2005). See below for PCL-6 score definition.

*Cases*

- Responded “a little bit”, “moderately”, “quite a bit” or “extremely” to any of the following questions: “Next is a list of problems and complaints that people sometimes have in response to such extremely stressful experiences. Please indicate how much you have been bothered by that problem in the past month:”
  - “Repeated, disturbing memories, thoughts or images of a stressful experience?” (data-field 20497)
  - “Feeling very upset when something reminded you of a stressful experience?” (data-field 20498)
  - “Avoiding activities or situations because they reminded you of a stressful experience?” (data-field 20495)
- AND total PCL-6 symptom severity score ≥ 14

*Controls*

- Responded to data-fields 20487, 20498 and 20495
- AND total PCL-6 symptom severity score < 14

***Self-harm***

Self-harm cases and controls were defined by “yes” or “no” answers to the following question: “Have you ever deliberately harmed yourself, whether or not you meant to end your life?” (data-field 20480)

**References.**

Als, T. D., Kurki, M. I., Grove, J., Voloudakis, G., Therrien, K., Tasanko, E., . . . Børglum, A. D. (2023). Depression pathophysiology, risk prediction of recurrence and comorbid psychiatric disorders using genome-wide analyses. *Nat Med, 29*(7), 1832-1844. doi:10.1038/s41591-023-02352-1

Burns, A. C., Windred, D. P., Rutter, M. K., Olivier, P., Vetter, C., Saxena, R., . . . Cain, S. W. (2023). Day and night light exposure are associated with psychiatric disorders: an objective light study in >85,000 people. *Nature Mental Health, 1*(11), 853-862. doi:10.1038/s44220-023-00135-8

Carvalho, A. F., Takwoingi, Y., Sales, P. M. G., Soczynska, J. K., Köhler, C. A., Freitas, T. H., . . . Vieta, E. (2015). Screening for bipolar spectrum disorders: A comprehensive meta-analysis of accuracy studies. *Journal of affective disorders, 172*, 337-346. doi:<https://doi.org/10.1016/j.jad.2014.10.024>

Cerimele, J. M., Chwastiak, L. A., Dodson, S., & Katon, W. J. (2014). The prevalence of bipolar disorder in general primary care samples: a systematic review. *General Hospital Psychiatry, 36*(1), 19-25. doi:<https://doi.org/10.1016/j.genhosppsych.2013.09.008>

Dashti, H. S., Daghlas, I., Lane, J. M., Huang, Y., Udler, M. S., Wang, H., . . . andMe Research, T. (2021). Genetic determinants of daytime napping and effects on cardiometabolic health. *Nature Communications, 12*(1), 900. doi:10.1038/s41467-020-20585-3

Davis, K. A. S., Coleman, J. R. I., Adams, M., Allen, N., Breen, G., Cullen, B., . . . Hotopf, M. (2020). Mental health in UK Biobank - development, implementation and results from an online questionnaire completed by 157 366 participants: a reanalysis. *BJPsych Open, 6*(2), e18. doi:10.1192/bjo.2019.100

Docherty, A. R., Mullins, N., Ashley-Koch, A. E., Qin, X., Coleman, J. R., Shabalin, A., . . . Adams, M. (2023). GWAS meta-analysis of suicide attempt: identification of 12 genome-wide significant loci and implication of genetic risks for specific health factors. *American Journal of Psychiatry, 180*(10), 723-738.

Doherty, A., Jackson, D., Hammerla, N., Plötz, T., Olivier, P., Granat, M. H., . . . Wareham, N. J. (2017). Large Scale Population Assessment of Physical Activity Using Wrist Worn Accelerometers: The UK Biobank Study. *PLOS ONE, 12*(2), e0169649. doi:10.1371/journal.pone.0169649

Friligkou, E., Løkhammer, S., Cabrera-Mendoza, B., Shen, J., He, J., Deiana, G., . . . Polimanti, R. (2024). Gene Discovery and Biological Insights into Anxiety Disorders from a Large-Scale Multi-Ancestry Genome-wide Association Study. *Nature Genetics (in press)*, 2024.2002.2014.24302836. doi:10.1101/2024.02.14.24302836

Horne, J. A., & Ostberg, O. (1976). A self-assessment questionnaire to determine morningness-eveningness in human circadian rhythms. *International journal of chronobiology, 4*(2), 97-110. Retrieved from <http://europepmc.org/abstract/MED/1027738>

Jones, S. E., Lane, J. M., Wood, A. R., van Hees, V., Tyrrell, J., Beaumont, R. N., . . . Weedon, M. N. (2019). Genome-wide association analyses of chronotype in 697,828 individuals provides insights into circadian rhythms. *Nature Communications, 10*(1), 343. doi:10.1038/s41467-018-08259-7

Jones, S. E., van Hees, V., Mazzotti, D. R., Marques-Vidal, P., Sabia, S., van der Spek, A., . . . Wood, A. R. (2019). Genetic studies of accelerometer-based sleep measures yield new insights into human sleep behaviour. *Nature Communications, 10*(1), 1585. doi:10.1038/s41467-019-09576-1

Kessler, R. C., Andrews, G., Mroczek, D., Ustun, B., & Wittchen, H.-U. (1998). The World Health Organization Composite International Diagnostic Interview short-form (CIDI-SF). *International Journal of Methods in Psychiatric Research, 7*(4), 171-185. doi:<https://doi.org/10.1002/mpr.47>

Koenig, Z., Yohannes, M. T., Nkambule, L. L., Zhao, X., Goodrich, J. K., Kim, H. A., . . . Sahakian, N. (2024). A harmonized public resource of deeply sequenced diverse human genomes. *Genome Research*.

Lane, J. M., Jones, S. E., Dashti, H. S., Wood, A. R., Aragam, K. G., van Hees, V., . . . Bowden, J. (2019). Biological and clinical insights from genetics of insomnia symptoms. *Nature genetics, 51*(3), 387-393.

Lang, A. J., & Stein, M. B. (2005). An abbreviated PTSD checklist for use as a screening instrument in primary care. *Behaviour Research and Therapy, 43*(5), 585-594. doi:<https://doi.org/10.1016/j.brat.2004.04.005>

Manichaikul, A., Mychaleckyj, J. C., Rich, S. S., Daly, K., Sale, M., & Chen, W.-M. (2010). Robust relationship inference in genome-wide association studies. *Bioinformatics, 26*(22), 2867-2873.

Mullins, N., Forstner, A. J., O’Connell, K. S., Coombes, B., Coleman, J. R. I., Qiao, Z., . . . Psychiatry, H. A.-I. (2021). Genome-wide association study of more than 40,000 bipolar disorder cases provides new insights into the underlying biology. *Nature genetics, 53*(6), 817-829. doi:10.1038/s41588-021-00857-4

Nievergelt, C. M., Maihofer, A. X., Klengel, T., Atkinson, E. G., Chen, C.-Y., Choi, K. W., . . . Koenen, K. C. (2019). International meta-analysis of PTSD genome-wide association studies identifies sex- and ancestry-specific genetic risk loci. *Nature Communications, 10*(1), 4558. doi:10.1038/s41467-019-12576-w

Nuevo, R., Chatterji, S., Verdes, E., Naidoo, N., Arango, C., & Ayuso-Mateos, J. L. (2010). The Continuum of Psychotic Symptoms in the General Population: A Cross-national Study. *Schizophrenia Bulletin, 38*(3), 475-485. doi:10.1093/schbul/sbq099

Patterson, N., Price, A. L., & Reich, D. (2006). Population structure and eigenanalysis. *PLOS Genetics, 2*(12), e190.

Pedregosa, F., Varoquaux, G., Gramfort, A., Michel, V., Thirion, B., Grisel, O., . . . Dubourg, V. (2011). Scikit-learn: Machine learning in Python. *the Journal of machine Learning research, 12*, 2825-2830.

Phillips, A. J. K., Clerx, W. M., O’Brien, C. S., Sano, A., Barger, L. K., Picard, R. W., . . . Czeisler, C. A. (2017). Irregular sleep/wake patterns are associated with poorer academic performance and delayed circadian and sleep/wake timing. *Scientific Reports, 7*(1), 3216. doi:10.1038/s41598-017-03171-4

Purcell, S., Neale, B., Todd-Brown, K., Thomas, L., Ferreira, M. A., Bender, D., . . . Daly, M. J. (2007). PLINK: a tool set for whole-genome association and population-based linkage analyses. *The American journal of human genetics, 81*(3), 559-575.

Torsvall, L., & Åkerstedt, T. (1980). A diurnal type scale: construction, consistency and validation in shift work. *Scandinavian journal of work, environment & health*, 283-290.

Trubetskoy, V., Pardiñas, A. F., Qi, T., Panagiotaropoulou, G., Awasthi, S., Bigdeli, T. B., . . . Bertolino, A. (2022). Mapping genomic loci implicates genes and synaptic biology in schizophrenia. *Nature, 604*(7906), 502-508. doi:10.1038/s41586-022-04434-5

van Hees, V., Gorzelniak, L., Dean León, E. C., Eder, M., Pias, M., Taherian, S., . . . Brage, S. (2013). Separating Movement and Gravity Components in an Acceleration Signal and Implications for the Assessment of Human Daily Physical Activity. *PLOS ONE, 8*(4), e61691. doi:10.1371/journal.pone.0061691

van Hees, V., Sabia, S., Jones, S. E., Wood, A. R., Anderson, K. N., Kivimäki, M., . . . Trenell, M. (2018). Estimating sleep parameters using an accelerometer without sleep diary. *Scientific Reports, 8*(1), 1-11.

Wang, H., Lane, J. M., Jones, S. E., Dashti, H. S., Ollila, H. M., Wood, A. R., . . . Kantojärvi, K. (2019). Genome-wide association analysis of self-reported daytime sleepiness identifies 42 loci that suggest biological subtypes. *Nature Communications, 10*(1), 3503.

Windred, D. P., Jones, S. E., Russell, A., Burns, A. C., Chan, P., Weedon, M. N., . . . Phillips, A. J. K. (2021). Objective assessment of sleep regularity in 60 000 UK Biobank participants using an open-source package. *Sleep, 44*(12). doi:10.1093/sleep/zsab254
